# Symptoms of anxiety and depression in relation to work patterns during the first wave of the COVID-19 epidemic in Philadelphia PA: a cross-sectional survey

**DOI:** 10.1101/2021.01.21.21250117

**Authors:** Igor Burstyn, Tran Huynh

## Abstract

**Objective:** We investigated whether patterns of work during COVID-19 pandemic altered by effort to contain the outbreak affected anxiety and depression.

**Methods:** We conducted a cross-sectional online survey of 911 residents of Philadelphia, inquiring about their working lives during early months of the epidemic, symptoms of anxiety and depression, plus demographics, perceived sources of support, and general health.

**Results:** Occupational contact with suspected COVID-19 cases was associated with anxiety. Concerns about return to work, childcare, lack of sick leave, and loss/reduction in work correlated with anxiety and depression, even when there was no evidence of occupational contact with infected persons; patterns differed by gender.

**Conclusions:** Heightened anxiety and depression during COVID-19 pandemic can be due to widespread disruption of working lives, especially in “non-essential” low-income industries, on par with experience in healthcare.

The significance to clinical practice of the information being presented: *Anxiety and depression symptoms that emerged during COVID-19 pandemic may be related to disruption of working lives even among people who are not the “essential” workers with one-one-one contact with infected persons. Clinicians may find this evidence of occupational correlates and articulated specific worries useful in treating such patients*.

## Introduction

The mental health impacts of the coronavirus disease (COVID-19) pandemic in the United States was evaluated by Czeisler et al.,(1) providing evidence of increase in anxiety and depression during April-June 2020 compared to the same period a year before, with a notable excess of “essential” workers having considered suicide. A nation-wide convenience sample (high in emergency department staff) of 2,040 healthcare workers during May 2020 revealed that having reported symptoms consistent with COVID-19 was associated with anxiety and depression.(2) Almost a third of the participants were suspected of having COVID-19, limiting works’ generalizability due to far lower prevalence of the disease even among healthcare workers at that time. First responders from Rocky Mountain region of the US during spring of 2020,(3) exhibited evidence of excess of anxiety and depression associated with reported contact with COVID-19 patients. A Canadian survey of mostly unionized professions outside of healthcare conducted during the first wave of pandemic, reported elevated rates of anxiety and depression, especially among those who could not work remotely (telecommute) or lost work; among those who had to have one-on-one contact with people at work, anxiety and depression was more common when their expectations of infection control were not met.(4) There are limited data on the contribution of work disruption by COVID-19 pandemic on mental health in the US, outside of healthcare workers and first responders. It is reasonable to suppose that effective (and perceived as such) measures that protect population from infectious disease outbreak will lessen anxiety related to the outbreak and may dampen any potential increase in mood disorders. However, such measures may have unintended negative consequences on mental health though disruption of economic activities and routines followed by families, e.g., those who rely on childcare (either via pre-schools or secondary schools) and with living arrangements not conducive to work from home/telecommuting.

The first case of COVID-19 in Philadelphia was announced on March 10, 2020. The city’s mayor issued the stay-at-home order on March 23,(3, 5) limiting business operations to “life-sustaining” and encouraging teleworking when feasible. The order targeted whole industries, not specific occupations. The stay-at-home was lifted on June 5 when Philadelphia could move from the “red” (most restrictive) to the “yellow” (less restrictive) phase of the Pennsylvania’s reopening plan. During the yellow phase, some essential businesses such as childcare centres could be open at limited capacity. The city could move to have even fewer restrictions on June 26 but kept them in place until July 3, when all businesses could reopen at 50% occupancy. Following these restrictions, Philadelphia experienced the highest unemployment of 2020 in July (135,295 claims), more than doubling since March (51, 297) and almost tripled compared to previous June (45,606).(6) Persons who already had low income and no chance to telecommute due to work in arts and entertainment, retail, and food industries were most adversely affected. Work is a main source of financial stability for many families, which in turn is related to mental health, with evidence that unemployment independently causes anxiety and depression, more so in men than women.(7) Therefore, considerations of changes to work, including unemployment on mental health, seems essential to ensuring that measures taken to combat infections outbreaks minimize concomitant adverse effects on mental health.

Our aim is to describe symptoms of anxiety and depression in a sample of general population of Philadelphia, PA, in relation to features of work during COVID-19 epidemic, with emphasis on associations with perceived and actual changes in work precipitated by the outbreak, while accounting for sources of support and general health.

## Methods

### Survey instrument

The survey contained blocks of questions that covered general health, demographics, work-related questions, perceptions, worries, and concerns about the pandemic, COVID-19 testing, and Hospital Anxiety and Depression Scale (HADS).(8, 9) HADS contains sub-scales that measure anxiety and depression separately, each ranging from 0-21, with scores ≥11 commonly used to identify cases in the general adult population. Two general perceived health questions from the SF-36 (36-Item Short Form Health Survey) were asked: (a) “in general, would you say your health is: excellent, very good, good, fair, poor” and “compared to other persons your age, would you say your health is: excellent, very good, good, fair, poor.”(10) Survey included a battery of questions about perceptions (captured on Likert-like scale ranging from 0 to 100) of working conditions in the most recent week of work, source of anticipated support during pandemic, and specific worries. “Worrying” is an established proximal antecedent of generalized anxiety (such as assessed by HADS) as opposed to a more distal “environmental” cause. (11, 12) Consequently, we did not adjust for worries in regression models of HADS scores described below, but rather (a) investigated association between worries and HADS for anxiety in principal components analysis and (b) used reported worries descriptively with respect to their correlation with HADS scores. Copies of research instruments are available upon request, but the key questions not present in the cited literature are reported as part of results below. The participants could choose to complete the survey in English, Spanish, Vietnamese, or Chinese.

### Recruitment

Eligible participants were adults aged 18 years and older and were living in Philadelphia, Pennsylvania. The study was restricted to Philadelphia residents because the lockdown date and policies mitigating the pandemic differed by county. Our data collection started on April 17 and ended July 3, 2020, spanning both red and yellow phases of restrictions.

The online survey was administered via Qualtrics software (Qualtrics, Provo, UT). Participants were recruited using a convenience sampling approach via multiple communications strategies. Emails were sent to the investigators’ network (heavily weighted to academic community), 623 registered community organizations using a publicly available roster (three mailings, between April 21 and May 13), and other community groups found on the internet. The survey was advertised in a neighbourhood online newspaper *West Philly Local* (on April 18)(13) and a regional newspaper’s website the *Philadelphia Inquirer* (May 11-17). Starting May 19, we used Facebook to design and customize an advertisement campaign to place ads on its site and affiliated social media platforms (Instagram, Messenger, and the Facebook Audience Network), following methodology in Ali et al.(14) Among these recruitment strategies, email distributions and social media appeared to yield the most responses.

### Data preparation

A total of 2,664 persons read the informed consent page and provided a response, of whom 1,577 consented to proceed with the survey. We discarded 283 participants whose HADS responses were missing more than 50% in each subscale as recommended by Bell et al.(15) We also removed 20 participants who indicated their gender were either “other” or missing due to the need to conduct stratified analysis by gender. The resulting number of subjects after these restrictions was 1,280. For this paper, we focused on the 911 participants who had a job since the first case of COVID-19 was reported in Philadelphia.

Participants provided free text responses about their jobs and what their employers do or make, that were then recorded by the authors into more interpretable occupation and industry categories. The entire coding scheme is captured in **Supplemental Material** 1.

There were missing values in most categorical and continuous variables. HADS scores with less than half of missing values were imputed with the individual subscale mean score.(15) Similarly, missing values in other continuous variables were imputed with means of observed values. Missing values of categorical variables were kept as is to stabilize regression analyses and more fully utilize the data.

### Statistical analysis

Data was prepared for analysis in R.(16) All statistical calculations were performed in SAS v 9.4 (SAS Institute, Cary, NC). Association of HADS scores for anxiety (HADS A) and depression (HADS D) were examined for each of the covariate of interest in terms of counts of scores ≥11 (referred to as “cases’’ hereafter) for categorical covariates and Spearman rank correlations for continuous covariates. Univariate associations of continuous HADS scores with categorical variables were evaluated in Kruskal-Wallis (K-W) tests. All analyses were stratified by gender due to known differences in (a) rates of anxiety and depression by sex and (b) working conditions between men and women even when the description of work appears identical. Multivariable regression models of HADS scores were estimated using binomial regression (PROC GENMOD). These yielded relative rates (RR) and 95% confidence intervals (CI) of change in HADS scores in relation to variables that showed evidence of association with HADS scores in univariate analyses. Regression analyses examined impact of industries and occupations employed effect coding such that the effect estimates are in relation to unweighted sample average. We assumed the following causal pathway and did not adjust effects of industry for specific work characteristics as they lay on the path towards the outcomes: stay-at-home-order → industry → specific work characteristics → anxiety/depression.

## Results

### Persons who reported a positive test for COVID-19

Fourteen participants reported that they had a “reason to believe that” they “may have been infected with the COVID-19 virus”; they all reported that they were unwell at least for two consecutive days since start of the epidemic in the city, and they reported to have tested positive for having been “infected with COVID-19 virus”. All identified with “white” race, four were men; all but one were 35 years of age or older and six were 55 years of age or older. Eight (57%) had HADS score for anxiety that qualified them as a case (≥11) and three (21%) were cases of depression (i.e., HADS depression score ≥11). Anxiety HADS scores (mean=median: 11, standard deviation (SD): 4.4, range 4-17) were substantially higher than depression ones (mean: 6.8, median=4.0, SD: 4.0, range 0-13), but closely related (correlation 0.8). Eight reported to have used PPE at work since start of the epidemic and one did not respond (8/13=62%), and eight reported to that “since March 10^th^” their “work involved one-on-one contact with known or suspected people with COVID-19” but 13 did not (62%). There was only moderate agreement between PPE use at work and work-related contact with known or suspected COVID-19 cases (correlation 0.4). All physicians and nurses (5) except one, and a paralegal reported to have used PPE when they knew they had contact with infected patients (6/8=75%) at work. A lawyer and a nurse did not use PPE when reportedly exposed to COVID-19 cases through work. Infected persons with no reported exposure to cases of COVID-19 were employees of a community non-profit association, attorneys, public relations professionals, and massage therapists. The other settings where infected participants worked included hospitals (including intensive care units), geriatric/nursing home facilities, and law firms. Due to small number of persons who reported to have tested positive for COVID-19, we excluded them from subsequent statistical analysis and no further examination of associations within that group were attempted.

### Persons who did not report a positive test for COVID-19

Detailed description of continuous HADS scores by categories presented in Tables **1a** and **1b** are in **Supplemental Material** 2.

**Table 1a:**
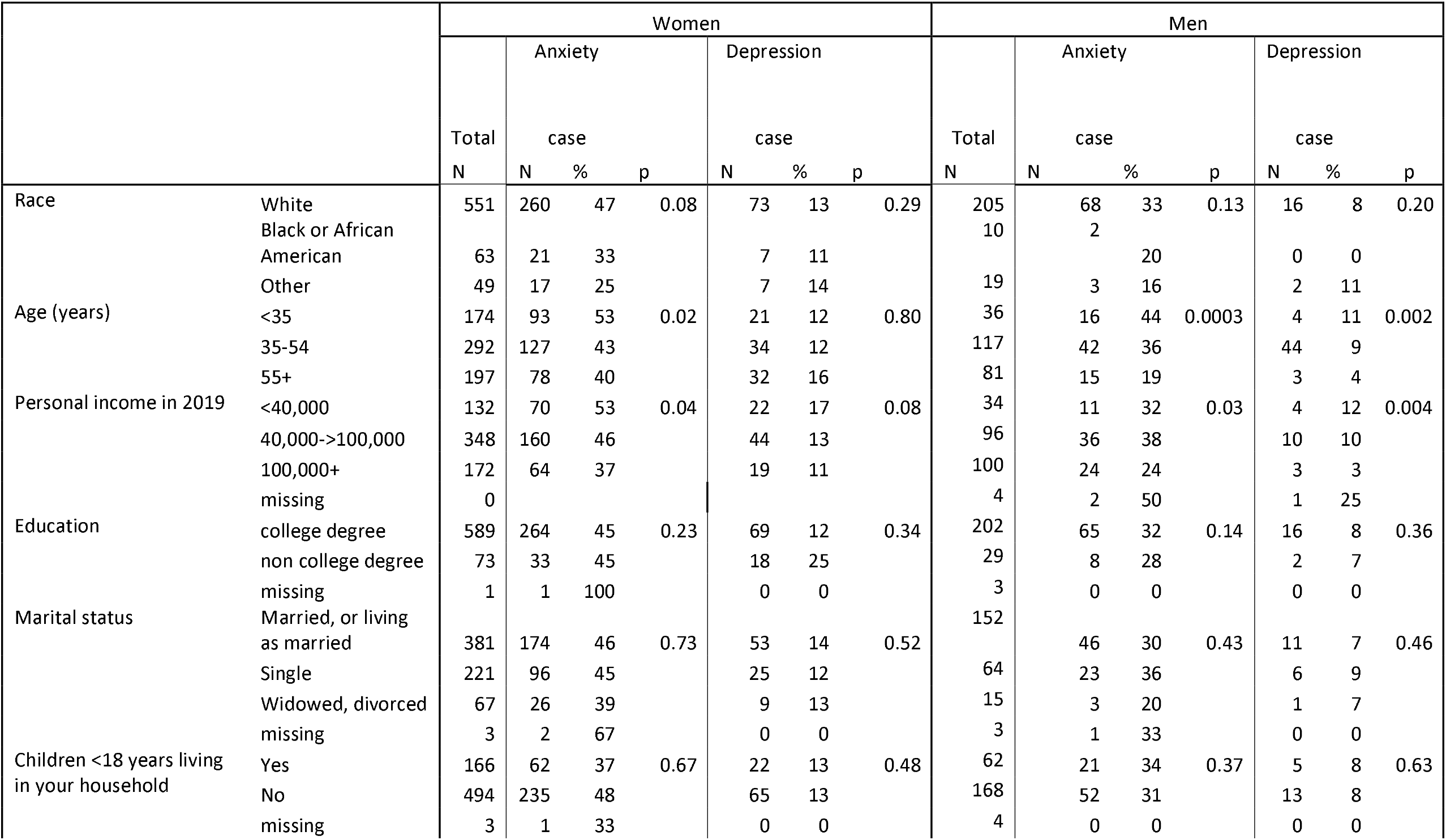
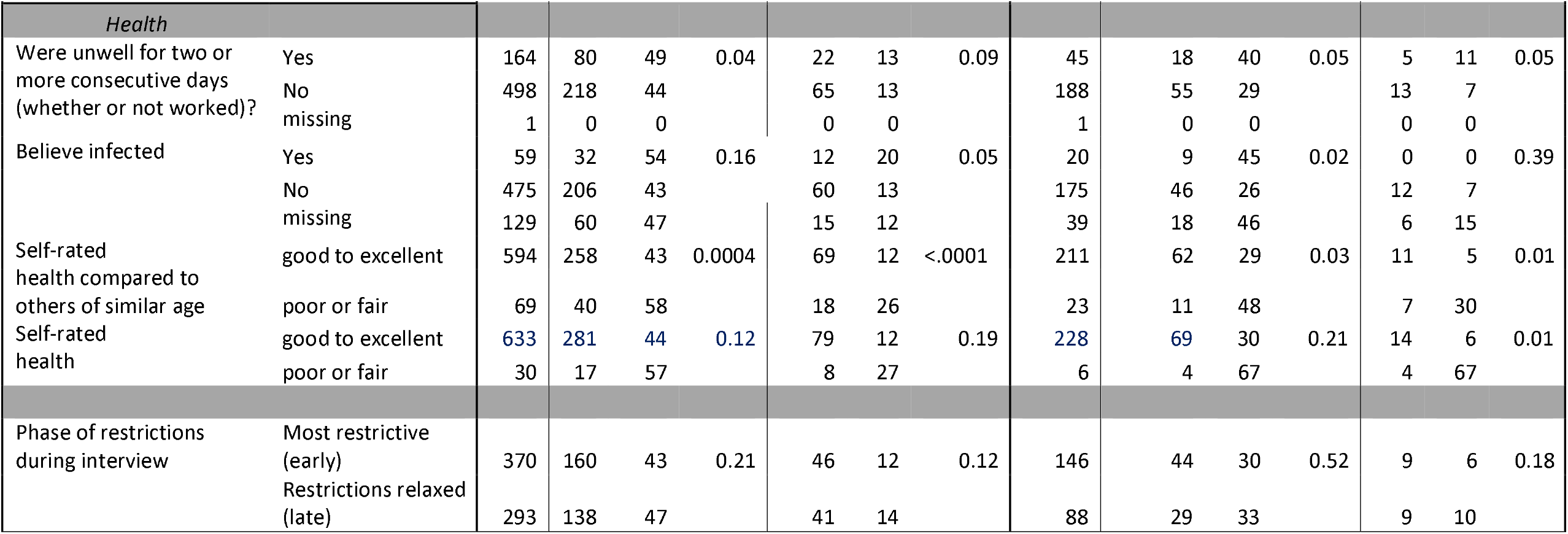
Demographic and health factors in relation to rates of anxiety and depression (HADS scores≥11); p-value is for Kruskal-Wallis test for difference in continuous HADS scores

**Table 1b:** Work-related factors in relation to rates of anxiety and depression (HADS scores≥11) among all who had a job (top) and those who reported change in work during since start of epidemic (bottom); p-value is for Kruskal-Wallis test for difference in continuous HADS scores

|  |  | Women |  |  |  |  |  |  | Men |  |  |  |  |  |  |
| --- | --- | --- | --- | --- | --- | --- | --- | --- | --- | --- | --- | --- | --- | --- | --- |
|  |  | Total<br>N | Anxiety |  |  | Depression |  |  | Total<br>N | Anxiety |  |  | Depression |  |  |
|  |  |  | case<br>N | % | p | case<br>N | % | p |  | case<br>N | % | p | case<br>N | % | p |
| *At work since start of epidemic* |  |  |  |  |  |  |  |  |  |  |  |  |  |  |  |
| one-on-one contact with people | Yes | 350 | 157 | 45 | 0.44 | 35 | 10 | 0.02 | 133 | 44 | 33 | 0.24 | 11 | 8 | 0.42 |
|  | No | 313 | 141 | 45 |  | 52 | 17 |  | 101 | 29 | 29 |  | 7 | 7 |  |
| one-on-one contact with known or suspected people with Covid-19 | Yes | 48 | 20 | 42 | 0.94 | 3 | 6 | 0.82 | 20 | 8 | 40 | 0.33 | 1 | 5 | 0.97 |
|  | No | 513 | 230 | 45 |  | 70 | 14 |  | 181 | 51 | 28 |  | 14 | 8 |  |
|  | Do not know | 102 | 48 | 47 |  | 14 | 14 |  | 33 | 14 | 42 |  | 3 | 9 |  |
| If there was contact, did you use PPE? | Yes | 40 | 15 | 38 | 0.08 | 0 | 0 | 0.002 | 16 | 6 | 38 | 0.63 | 0 | 0 | 0.27 |
|  | No | 8 | 5 | 63 |  | 3 | 38 |  | 4 | 2 | 50 |  | 1 | 25 |  |
| Employment status at time of survey | salaried | 433 | 199 | 46 | 0.83 | 49 | 11 | 0.25 | 155 | 45 | 29 | 0.83 | 9 | 6 | 0.33 |
|  | hourly | 124 | 56 | 45 |  | 20 | 16 |  | 36 | 13 | 39 |  | 2 | 6 |  |
|  | contractor | 67 | 29 | 43 |  | 12 | 17 |  | 34 | 11 | 32 |  | 6 | 18 |  |
|  | other | 34 | 11 | 32 |  | 4 | 12 |  | 9 | 4 | 44 |  | 1 | 11 |  |
|  | missing | 5 | 3 | 60 |  | 2 | 40 |  | 0 |  |  |  | 0 |  |  |
| Days a week worked | 5+ | 497 | 231 | 46 | 0.20 | 61 | 12 | 0.87 | 190 | 64 | 34 | 0.25 | 14 | 7 | 0.90 |
|  | <5 | 166 | 67 | 40 |  | 26 | 16 |  | 44 | 9 | 20 |  | 4 | 9 |  |
| Hours a week worked | 40+ | 365 | 169 | 46 | 0.29 | 50 | 14 | 0.37 | 154 | 51 | 33 | 0.25 | 11 | 7 | 0.83 |
|  | <40 | 298 | 129 | 43 |  | 37 | 12 |  | 80 | 22 | 28 |  | 7 | 9 |  |
| Access to paid sick leave/disability through work | Yes | 510 | 234 | 46 | 0.37 | 59 | 12 | 0.47 | 172 | 50 | 29 | 0.81 | 9 | 5 | 0.14 |
|  | No | 133 | 53 | 40 |  | 23 | 17 |  | 57 | 22 | 39 |  | 9 | 16 |  |
|  | Unsure | 20 | 11 | 55 |  | 5 | 25 |  | 5 | 1 | 20 |  | 0 | 0 |  |
| Essential worker? | Yes | 126 | 51 | 41 | 0.59 | 12 | 10 | 0.43 | 70 | 23 | 33 | 0.56 | 5 | 7 | 0.36 |
|  | No | 485 | 222 | 46 |  | 67 | 14 |  | 151 | 45 | 30 |  | 12 | 8 |  |
|  | maybe | 52 | 25 | 48 |  | 8 | 15 |  | 13 | 5 | 38 |  | 1 | 8 |  |
|  |  | Total<br>N | Anxiety |  |  | Depression |  |  | Total<br>N | Anxiety |  |  | Depression |  |  |
|  |  |  | N | % | p | N | % | p |  | N | % | p | N | % | p |
| Period unable to work due to high risk of spreading of COVID-19 | Yes | 76 | 36 | 47 | 0.59 | 11 | 14 | 0.13 | 22 | 8 | 36 | 0.90 | 3 | 14 | 0.49 |
|  | No | 578 | 262 | 45 |  | 76 | 13 |  | 212 | 65 | 31 |  | 15 | 7 |  |
| Period unable to work for other reasons | Yes | 136 | 54 | 40 | 0.44 | 15 | 11 | 0.34 | 40 | 16 | 30 | 0.65 | 2 | 5 | 0.57 |
|  | No | 527 | 244 | 46 |  | 72 | 14 |  | 193 | 57 | 40 |  | 16 | 8 |  |
| *Work arrangement changes due to epidemic* |  |  |  |  |  |  |  |  |  |  |  |  |  |  |  |
| Work arrangement changed by Stay-at-home order of March 23 | Yes | 489 | 225 | 46 | 0.61 | 62 | 13 | 0.94 | 155 | 54 | 35 | 0.03 | 12 | 8 | 0.37 |
|  | No | 173 | 72 | 42 |  | 25 | 14 |  | 79 | 19 | 24 |  | 6 | 8 |  |
|  | missing | 1 | 1 | 1 |  | 0 | 0 |  | 0 |  |  |  | 0 |  |  |
| Lost job | & applied unemployment | 38 | 18 | 47 | 0.40 | 4 | 11 | 0.17 | 10 | 6 | 60 | 0.52 | 1 | 10 | 0.03 |
|  | & did not apply unemployment | 14 | 7 | 50 |  | 4 | 29 |  | 4 | 4 | 100 |  | 2 | 50 |  |
|  | & missing | 1 | 0 | 0 |  | 0 | 0 |  | 0 |  |  |  |  |  |  |
|  | No | 436 | 200 | 46 | 0.87* | 54 | 12 | 0.52* | 141 | 44 | 31 | 0.05* | 9 | 6 | 0.007* |
| Work hours change if not lost job | Same | 214 | 93 | 43 | 0.02 | 25 | 12 | 0.23 | 69 | 19 | 28 | 0.53 | 2 | 3 | 0.24 |
|  | More | 138 | 56 | 41 |  | 20 | 14 |  | 42 | 14 | 33 |  | 6 | 14 |  |
|  | Fewer | 83 | 50 | 60 |  | 9 | 11 |  | 30 | 11 | 37 |  | 1 | 3 |  |
|  | missing | 1 | 1 | 100 |  | 0 | 0 |  | 0 |  |  |  |  |  |  |
| Struggle with balancing work and childcare, if kids <18 at home | Yes | 53 | 19 | 36 | 0.99 | 5 | 9 | 0.49 | 16 | 9 | 56 | 0.29 | 2 | 13 | 0.11 |
|  | No | 55 | 21 | 38 |  | 8 | 15 |  | 25 | 6 | 24 |  | 1 | 4 |  |
|  | missing | 19 | 7 | 37 |  | 2 | 11 |  | 8 | 3 | 38 |  | 0 | 0 |  |
| Start or substantially increase telecommuting | Yes | 342 | 155 | 45 | 0.78 | 42 | 12 | 0.91 | 105 | 36 | 34 | 0.91 | 6 | 6 | 0.003 |
|  | No | 55 | 27 | 49 |  | 7 | 13 |  | 18 | 5 | 28 |  | 1 | 6 |  |
|  | missing | 92 | 43 | 48 |  | 13 | 14 |  | 32 | 13 | 41 |  | 5 | 17 |  |
| Concerned about return to work |  | 336 | 171 | 51 | 0.0003 | 49 | 15 | 0.007 | 88 | 39 | 44 | 0.008 | 9 | 10 | 0.006 |

\* p-values K-W test for lost job vs not.

Most participants in our survey who had a job at one time during the epidemic (663 women and 234 men) did not report that they tested positive for COVID-19 at the time of survey, although among them 59 women and 20 men believed that they have been infected. The demographics of person with no reported positive test for COVID-19 are presented in **Table 1a**, showing that they were predominantly white, aged 35-54 years, with personal income >$40,000 in 2019, completed college, were married, and did not have children under 18 years of age living at home. Clearly the participants do not represent typical residents of Philadelphia, a city that is far more diverse than our sample. For both men and women, there is evidence of increased rates of anxiety among those who identify as white, are <35 years of age, and with personal income <$40,000 in 2019 (rates of anxiety about 50% in women and 30-40% in men). There is evidence of excess of depression among women and men with personal income <$40,000 in 2019, and among younger men (rates approaching 20% in women and approximately 10% in men).

Among women, 298 (45%) had HADS score for anxiety that qualified them as a case, and 87 (13%) were cases of depression. Among men, 73 (31%) had HADS score for anxiety that qualified them as a case, and 18 (8%) were cases of depression, both lower than among women. These patterns are corroborated by distribution of continuous HADS scores. Among women, anxiety HADS scores (mean=median: 10, SD: 4.1, range 0-21) were worse than among men (mean: 8.5, median: 8, SD: 4.2, range 0-20). Likewise, among women, depression HADS scores (mean: 6.5, median: 6, SD: 3.7, range 0-20) were worse than among men (mean: 5.4, median: 5, SD: 3.5, range 0-18). The rank correlation of depression and anxiety scores was 0.6 (p<0.0001).

Among persons who did not report a positive test for COVID-19, 25% of women and 19% of men reported to have been unwell for two or more consecutive days since start of the epidemic. This is the group that appeared to have elevated rates and HADS scores for both anxiety and depression, e.g., rates of anxiety among women of 49%, and among men -- 40%, p=0.04 and p=0.05 for K-W tests on continuous scale of HADS, respectively. There were similar associations among those who believe they were infected: 9% of women and men (though most were untested). “Poor to fair” self-rated health, especially in comparison to “others of the same age” was linked to higher rates of anxiety and depression across genders. The phase of restrictions aimed to mitigate the epidemic during which data was collected did not appear to have an effect.

The distribution of work-related factors and their associations with anxiety and depression are described in **Table 1b**. Top part of the table relates to all who reported having had a job during the epidemic and the bottom part – to a subset who reported change in work since start of the epidemic in Philadelphia. About half of men and women reported to have had one-on-one contact with people at work since start of the epidemic, with women more likely to be depressed if they had no such contact (52 case, 17% rate vs. 10%). There was no evidence of associations of contact with known or suspected people with COVID-19 through work with mood disorder unless such contact occurred without the protection of PPE. The number of such cases is admittedly small, but trend is consistent across disorders and genders, being stronger for in women: for anxiety, based on 5 cases crude rate of 63% (p=0.08 on continuous scale), and for depression, based 3 cases crude rate of 38% (p=0.002 on continuous scale). With respect to type of employment arrangement, the highest rates of anxiety were among hourly employees and depression – among contractors, but these patterns may well be due to chance alone. Working longer hours appeared to be associated with higher anxiety, albeit weakly (p≤0.3, 3-6% difference), but not depression. Lack of access to sick leave or disability through work was associated with elevated depression rate in men (16 vs 5%; p=0.14) and suggestion of a similar effect in women (17 vs 12%; but p=0.47). Persons who reported to be essential workers had the same rates of outcomes as those who did not. Report of not having been able to work for a period due to high risk of spread of COVID-19 (but not other reasons) was related to higher rate of depression only in women (14 vs 13%, p=0.13), although crude rates suggest this effect for all outcomes and genders.

Most participants in our survey (489 women, 74%, and 155 men, 66%) reported that their work arrangements changed due to epidemic (or at least since its start). Only in men did this change relate to elevated anxiety (35 vs 24%; p=0.03). Having lost a job was associated with elevated rates of anxiety and depression in men only. Although the numbers are small, those who did not yet apply for unemployment benefits appeared more anxious and depressed than those who did, across both genders. Women who reported having reduced working hours were the only ones who were more anxious (50 cases, crude rate 60%) but not depressed. In men, but not women, there was evidence of elevated anxiety and depression in conjunction with both having kids live at home and struggling to balance work and childcare, albeit numbers are small. There was an indication that start or increase in in remote work was linked with depression in men but only based on difference in HADS scores (p=0.003), not rate of cases. There was robust evidence of “concerned about returning to work immediately after stay-at-home order is lifted in the future” being related to elevated rates of anxiety and depression across genders (all p<0.01); this concern was reported by 336/489=69% women and 88/155=57% men. Specific concerns mentioned in free text form by 425 respondents were overwhelmingly (370, 87%) only about risk of infection, including at work and on the way to work (via public transit). The rest (55, 13%) predominantly dealt with concerns about availability of childcare and there being no jobs to return to. These univariate associations were jointly considered in regression analyses presented below, after presentation of description of perceptions, supports, and worries.

The reported perceptions of work, supports, and worries are summarized in **Table 2** for all participants who were employed since epidemic started in Philadelphia. On average, participants neither agreed nor disagreed that their work hours and tasks changed during the most recent week, with average scores of 50 across genders (on a scale of 0 to 100). These perceptions were not related to anxiety and depression in rank corelations. The dominant source of perceived support during the epidemic was “immediate family”, with average scores 80 out of 100. Believing that one will find support within immediate family was the strongest consistent correlate of lower anxiety and depression scores across genders. Co-workers, employers, personal physician, and neighbours were the second strongest sources of reported support, with scores 40-50. Federal government, social service organizations and trade unions were the least commonly reported sources of support, with scores in 20s and below. There was difference between men and women in correlation of these supports with anxiety and depression scores. Among men, in addition to the protective effect of strong family support, only having a supportive employer was related to fewer symptoms of anxiety and depression. Among women, in addition to supportive employer, perceived support from co-workers, federal government, and religious community was related to fewer symptoms of anxiety and depression (correlations −0.1 to −0.2, p<0.002).

**Table 2:** Anxiety and depression scores in relations to reported perceptions, support and worries; Anx=anxiety; Dep=depression

|  | Women (n=623) |  |  |  |  |  |  | Men (n=234) |  |  |  |  |  |  |
| --- | --- | --- | --- | --- | --- | --- | --- | --- | --- | --- | --- | --- | --- | --- |
|  | Mean | SD | Median | rank correlation (p-value) |  |  |  | Mean | SD | Median | rank correlation (p-value) |  |  |  |
|  |  |  |  | HADS Anx |  | HADS Dep |  |  |  |  | HADS Anx |  | HADS Dep |  |
| Perception of work during recent week of epidemic (completely disagree=0, completely agree=100) |  |  |  |  |  |  |  |  |  |  |  |  |  |  |
| My hours of work are about the same | 52 | 34 | 52 | -0.04 | 0.30 | -0.03 | 0.38 | 51 | 33 | 52 | 0.05 | 0.4 | 0.07 | 0.3 |
| My work tasks are about the same | 52 | 30 | 52 | 0.03 | 0.39 | -0.01 | 0.83 | 51 | 30 | 52 | 0.07 | 0.3 | 0.08 | 0.3 |
| Where you will find support (no support at all =0, very strong support=100) |  |  |  |  |  |  |  |  |  |  |  |  |  |  |
| My immediate family | 80 | 26 | 91 | -0.19 | <.0001 | -0.19 | <.0001 | 81 | 27 | 93 | -0.19 | 0.003 | -0.20 | 0.002 |
| My co-workers | 55 | 29 | 50 | -0.19 | <.0001 | -0.16 | <.0001 | 51 | 30 | 49 | -0.05 | 0.4 | -0.09 | 0.2 |
| My employer | 48 | 30 | 43 | -0.16 | <.0001 | -0.17 | <.0001 | 46 | 30 | 43 | -0.12 | 0.07 | -0.16 | 0.02 |
| My doctor | 43 | 29 | 45 | -0.12 | 0.002 | -0.12 | 0.002 | 43 | 27 | 45 | -0.06 | 0.3 | -0.09 | 0.2 |
| Federal Government | 22 | 20 | 23 | -0.12 | 0.001 | -0.12 | 0.002 | 26 | 22 | 23 | -0.08 | 0.2 | -0.05 | 0.4 |
| City of Philadelphia | 37 | 24 | 36 | -0.05 | 0.24 | -0.09 | 0.02 | 35 | 23 | 36 | 0.04 | 0.6 | 0.06 | 0.4 |
| Department of Public Health (City) | 39 | 25 | 39 | -0.06 | 0.14 | -0.1 | 0.007 | 40 | 25 | 39 | 0.02 | 0.7 | -0.03 | 0.6 |
| My religious community | 38 | 28 | 38 | -0.14 | 0.0004 | -0.17 | <.0001 | 38 | 28 | 38 | -0.02 | 0.8 | -0.005 | 0.9 |
| Social services organization | 25 | 22 | 26 | -0.07 | 0.09 | -0.08 | 0.03 | 26 | 22 | 26 | 0.06 | 0.4 | 0.03 | 0.6 |
| My neighbours | 42 | 27 | 42 | -0.07 | 0.08 | -0.06 | 0.1 | 41 | 25 | 42 | -0.05 | 0.4 | -0.10 | 0.1 |
| My worker union | 15 | 18 | 15 | -0.06 | 0.15 | -0.08 | 0.05 | 17 | 20 | 15 | 0.06 | 0.3 | 0.06 | 0.3 |
| Other | 65 | 20 | 64 | -0.04 | 0.31 | -0.02 | 0.7 | 63 | 18 | 64 | -0.04 | 0.5 | -0.18 | 0.01 |
| Worries about the COVID-19 epidemic (not at all worried=0, very worried=100) |  |  |  |  |  |  |  |  |  |  |  |  |  |  |
| I will be infected | 62 | 27 | 62 | 0.24 | <.0001 | 0.21 | <.0001 | 57 | 28 | 60 | 0.25 | 0.0001 | 0.18 | 0.01 |
| I will infect my family | 63 | 32 | 68 | 0.24 | <.0001 | 0.18 | <.0001 | 57 | 31 | 61 | 0.28 | <.0001 | 0.15 | 0.02 |
| I will not be able to cope with the work | 42 | 29 | 37 | 0.28 | <.0001 | 0.18 | <.0001 | 33 | 25 | 37 | 0.25 | 0.0001 | 0.25 | 0.00 |
| I will become poor | 42 | 31 | 40 | 0.25 | <.0001 | 0.20 | <.0001 | 38 | 27 | 40 | 0.35 | <.0001 | 0.30 | <.0001 |
| I will be short of food | 32 | 26 | 31 | 0.22 | <.0001 | 0.16 | <.0001 | 29 | 23 | 31 | 0.30 | <.0001 | 0.23 | 0.00 |
| I will be short of medicines | 31 | 26 | 31 | 0.17 | <.0001 | 0.12 | 0.002 | 29 | 23 | 31 | 0.25 | 0.0001 | 0.31 | <.0001 |
| I will fail myself and my family | 42 | 30 | 40 | 0.31 | <.0001 | 0.20 | <.0001 | 36 | 26 | 40 | 0.31 | <.0001 | 0.30 | <.0001 |
| I will be confined at home and not able to leave | 49 | 29 | 48 | 0.24 | <.0001 | 0.19 | <.0001 | 41 | 28 | 48 | 0.15 | 0.02 | 0.14 | 0.04 |

Among reported worries, those related to infecting oneself and one’s family dominated, with average scores around 60 out of 100 (very worried). The second most common variety of worry was that of being “confined at home and not able to leave”, with average scores 40-50 (higher among women). Being short on food and medical supplies was the least prominent worry, with average scores around

30. All worries were correlated with greater number of symptoms of anxiety and depression, as was confirm in principal component analysis (PCA) (**Supplemental Material 3**) that indicated that HADS anxiety scores and responses to worry questions all loaded onto one latent construct, with only one such construct dominant in explaining common variance (40%). PCA revealed two other principal components that are worth noting. The second captured worry about infection to self and family, but not worries regarding impoverishment and food shortages, and was barely related to anxiety (13-14% of variance). The third was gender-specific, capturing worries about confined to home, not being able to cope with work, and general anxiety among women, and worries about failing oneself and family together with general anxiety among men (10% of variance).

### Consideration of effect of changes in work

We next consider mutually adjusted effects work-related factors, demographics, general health, perceptions, and supports among the majority who reported changes in work since the first recorded case of COVID-19 in Philadelphia.

**Table 3** presents results of negative binomial *regression* of HADS anxiety score (on continuous scale) adjusted for all factors considered above; these results are not materially different when adjustment excluded perceptions and supports (details not shown). We did not have sufficient sample to obtain meaningful adjusted estimates of PPE use by suitable strata of known and suspected contact with infected persons, and PPE use per se was not associated with the outcome (not shown). Concern about return to work was the most consistent independent predictor of anxiety among work-related factors in both genders (women: RR 1.16, 95%CI: 1.07, 1.25; men: RR 1.23, 95% 1.06, 1.43). Working more than five days a week and having working hours reduced during epidemic appeared to be independently associated with increased anxiety in both genders. An extreme case of loss of working hours, losing a job, was the strongest correlate of anxiety in men, after allowing for other factors (RR1.56, 95%CI 1.12, 2.19). Perception of working hours being unchanged in the week before interview was related to reduced anxiety in women but increased anxiety in men. The more work tasks were perceived as having not changed in the week before interview, the more anxious women (but not men) were. Starting or substantially increasing telecommuting appeared to be associated with increased anxiety in both genders as well, with the effect more prominent among men. Men (but not women) who identified as essential workers (RR 1.16, 95%CI: 0.96, 1.40), had one-on one contact with people at work (RR 1.14, 95%CI: 0.98, 1.34), including known or suspected cases of COVID-19 (RR 1.30, 95%CI: 0.97, 1.74), who were hourly employees (RR 1.24, 95%CI: 0.96, 1.60), and did not have access to disability/sick leave through work (RR 1.22, 95%CI 0.93, 1.60) were more anxious. Reported struggle with balancing work and childcare was not independently related to anxiety after allowing for other factors. Only support from immediate family appeared protective across genders, with men additionally apparently benefitting from support from their unions. Curiously, participants of both genders who reported to be relying more heavily on city of Philadelphia for support showed more symptoms of anxiety. Among health-related factors, having been unwell with or without missing work was independently related to higher level of anxiety as was perception of poor general health compared to others of the same age (not shown).

**Table 3:** Effect on anxiety of work-related factors among those who reported change in work (489 women and 155 men): relative rate (RR) and 95% confidence intervals (CI) of the change in continuous HADS scores.

|  |  | Women |  |  | Men |  |  |
| --- | --- | --- | --- | --- | --- | --- | --- |
|  |  | RR* | 95%CI |  | RR* | 95%CI |  |
| work since start of epidemic |  |  |  |  |  |  |  |
| Essential worker? | yes | 0.97 | 0.87 | 1.09 | 1.16 | 0.96 | 1.40 |
|  | maybe | 1.03 | 0.91 | 1.16 | 0.89 | 0.66 | 1.20 |
|  | no | 1.00 |  |  | 1.00 |  |  |
| one-on-one contact with people |  | 0.97 | 0.90 | 1.04 | 1.14 | 0.98 | 1.34 |
| one-on-one contact with known or suspected people with Covid-19 | yes | 1.11 | 0.93 | 1.33 | 1.30 | 0.97 | 1.74 |
|  | do not know | 0.97 | 0.89 | 1.07 | 1.07 | 0.87 | 1.31 |
|  | no | 1.00 |  |  | 1.00 |  |  |
| Employment status at time of survey | contractor | 1.02 | 0.86 | 1.21 | 0.93 | 0.68 | 1.27 |
|  | hourly | 0.99 | 0.89 | 1.10 | 1.24 | 0.96 | 1.60 |
|  | other | 0.95 | 0.80 | 1.13 | 1.24 | 0.82 | 1.88 |
|  | salaried | 1.00 |  |  | 1.00 |  |  |
| Days a week worked | 5+ vs <5 | 1.05 | 0.96 | 1.15 | 1.14 | 0.93 | 1.39 |
| Hours a week worked | 40+ vs <40 | 1.02 | 0.94 | 1.10 | 1.05 | 0.89 | 1.24 |
| Access to paid sick leave/disability through work | no | 0.98 | 0.85 | 1.11 | 1.22 | 0.93 | 1.60 |
|  | unsure | 0.96 | 0.79 | 1.17 | 1.08 | 0.67 | 1.75 |
|  | yes | 1.00 |  |  | 1.00 |  |  |
| Period unable to work due to high risk of spreading of COVID-19 |  | 0.99 | 0.89 | 1.11 | 0.90 | 0.70 | 1.15 |
| Change in work hours | lost work | 0.94 | 0.78 | 1.13 | 1.56 | 1.12 | 2.19 |
|  | less | 1.11 | 1.00 | 1.23 | 1.09 | 0.89 | 1.34 |
|  | more | 0.97 | 0.88 | 1.06 | 0.86 | 0.68 | 1.09 |
|  | same | 1.00 |  |  | 1.00 |  |  |
| Did not start or substantially increase telecommuting |  | 1.03 | 0.92 | 1.16 | 1.21 | 0.96 | 1.54 |
| Concerned about return to work |  | 1.16 | 1.07 | 1.25 | 1.23 | 1.06 | 1.43 |
| Struggle with balancing work and childcare |  | 1.04 | 0.91 | 1.19 | 1.04 | 0.82 | 1.33 |
| perceptions of work in most recent week** |  |  |  |  |  |  |  |
| My hours of work are about the same |  | 0.97 | 0.93 | 1.00 | 1.04 | 0.97 | 1.12 |
| My work tasks are about the same |  | 1.03 | 1.00 | 1.07 | 0.96 | 0.90 | 1.02 |
| support** |  |  |  |  |  |  |  |
| My immediate family |  | 0.95 | 0.92 | 0.99 | 0.92 | 0.86 | 0.99 |
| My co-workers |  | 0.97 | 0.93 | 1.01 | 1.02 | 0.91 | 1.13 |
| My employer |  | 0.98 | 0.94 | 1.02 | 1.01 | 0.91 | 1.11 |
| City of Philadelphia |  | 1.05 | 0.99 | 1.11 | 1.08 | 0.95 | 1.22 |
| My worker union |  | 0.97 | 0.93 | 1.02 | 0.89 | 0.81 | 0.99 |
| Were unwell for two or more consecutive days (whether or not worked)? |  | 1.04 | 0.96 | 1.13 | 1.12 | 0.94 | 1.34 |

\* adjusted for age, education, income, children living at home, phase of stay-at-home order, general health; not all effect estimates for support shown; effect estimates for missing categories are not shown
\*\* estimates are per 25 units on Likert-like scale

**Table 4** presents results of negative binomial regression of HADS depression score (on continuous scale) adjusted for all factors considered in Table 3 in regression of HADS anxiety score as the outcome; these results are not materially different when adjustment excluded perceptions and supports (details not shown). As with anxiety, concern about return to work was the most consistent independent predictor of depression among work-related factors in both genders (women: RR 1.12, 95%CI: 1.00, 1.25; men: RR 1.26, 95% 1.04, 1.53). Working more than 40 hours a week (but not 5 days a week or more) appeared to be associated with increased depression in both genders, but the effect is most convincing among women (RR 1.13, 95%CI 1.00, 1.27). These effects were adjusted for perception of work hours and tasks in the most recent week, which on their own appeared unrelated to depression. Lack of access to disability or sick leave through work was associated with depression in both genders, but more so in men (RR 1.33, 95%CI 0.93, 1.90). Men (but not women) who lost a job were more depressed compared to those who worked same hours as before the epidemic, after allowing for other factors (RR1.25, 95%CI 0.82, 1.89). Unlike anxiety, the reported struggle with balancing work and childcare was independently related to depression after allowing for other factors, more so among men (RR 1.27, 95%CI: 0.93, 1.75) then women (RR 1.08, 95%CI 0.88, 1.31). Among both genders, hourly employees were more depressed relative to salaried ones. Having had one-on-one contact with people at work was related to fewer depression symptoms among women only (RR 0.91, 95%CI: 0.82, 1.01). Being an essential worker, contact with persons known or suspected to have COVID-19 at work, starting or substantially increasing telecommuting appeared to not be independently associated with depression in both genders. Both support from immediate family and trade unions were related to fewer symptoms of depression. As with anxiety, participants of both genders who reported to be relying more heavily on city of Philadelphia for support showed more symptoms of depression, especially men (RR 1.22, 95%CI 1.03, 1.44). Among health-related factors, having been unwell with or without missing work was independently related to higher level of depression as was perception of poor general health compared to others (not shown).

**Table 4:** Effect on depression of work-related factors among those who reported change in work (489 women and 155 men): relative rate (RR) and 95% confidence intervals (CI) of the change in continuous HADS scores.

|  |  | Women |  |  | Men |  |  |
| --- | --- | --- | --- | --- | --- | --- | --- |
|  |  | RR* | 95%CI |  | RR* | 95%CI |  |
| work since start of epidemic |  |  |  |  |  |  |  |
| Essential worker? | yes | 0.89 | 0.75 | 1.04 | 1.03 | 0.80 | 1.33 |
|  | maybe | 1.03 | 0.87 | 1.22 | 1.12 | 0.76 | 1.66 |
|  | no | 1.00 |  |  | 1.00 |  |  |
| one-on-one contact with people |  | 0.91 | 0.82 | 1.01 | 1.04 | 0.84 | 1.29 |
| one-on-one contact with known or suspected people with Covid-19 | yes | 1.08 | 0.83 | 1.41 | 1.10 | 0.74 | 1.64 |
|  | do not know | 0.94 | 0.82 | 1.08 | 0.99 | 0.75 | 1.30 |
|  | no | 1.00 |  |  | 1.00 |  |  |
| Employment status at time of survey | contractor | 1.05 | 0.83 | 1.33 | 0.96 | 0.64 | 1.44 |
|  | hourly | 1.09 | 0.93 | 1.27 | 1.26 | 0.90 | 1.77 |
|  | missing |  |  |  |  |  |  |
|  | other | 0.85 | 0.65 | 1.10 | 1.18 | 0.68 | 2.05 |
|  | salaried | 1.00 |  |  | 1.00 |  |  |
| Days a week worked | 5+ vs <5 | 0.97 | 0.85 | 1.11 | 1.00 | 0.78 | 1.29 |
| Hours a week worked | 40+ vs <40 | 1.13 | 1.00 | 1.27 | 1.08 | 0.86 | 1.34 |
| Access to paid sick leave/disability through work | no | 1.07 | 0.89 | 1.30 | 1.33 | 0.93 | 1.90 |
|  | unsure | 0.95 | 0.72 | 1.26 | 0.98 | 0.51 | 1.85 |
|  | yes | 1.00 |  |  | 1.00 |  |  |
| Period unable to work due to high risk of spreading of COVID-19 |  | 1.05 | 0.89 | 1.23 | 1.03 | 0.75 | 1.41 |
| Change in work hours | lost work | 1.03 | 0.79 | 1.34 | 1.25 | 0.82 | 1.89 |
|  | less | 1.09 | 0.93 | 1.27 | 1.04 | 0.79 | 1.37 |
|  | more | 1.05 | 0.92 | 1.20 | 1.02 | 0.75 | 1.39 |
|  | same | 1.00 |  |  | 1.00 |  |  |
| Did not start or substantially increase telecommuting |  | 0.95 | 0.81 | 1.12 | 1.16 | 0.85 | 1.56 |
| Concerned about return to work |  | 1.12 | 1.00 | 1.25 | 1.26 | 1.04 | 1.53 |
| Struggle with balancing work and childcare |  | 1.08 | 0.88 | 1.31 | 1.27 | 0.93 | 1.74 |
| perceptions of work in most recent week** |  |  |  |  |  |  |  |
| My hours of work are about the same |  | 0.99 | 0.94 | 1.04 | 0.99 | 0.90 | 1.09 |
| My work tasks are about the same |  | 1.00 | 0.95 | 1.06 | 0.99 | 0.91 | 1.08 |
| support** |  |  |  |  |  |  |  |
| My immediate family |  | 0.92 | 0.88 | 0.97 | 0.95 | 0.87 | 1.05 |
| My co-workers |  | 1.00 | 0.94 | 1.06 | 1.05 | 0.91 | 1.21 |
| My employer |  | 0.96 | 0.91 | 1.02 | 1.00 | 0.88 | 1.13 |
| City of Philadelphia |  | 1.04 | 0.96 | 1.12 | 1.22 | 1.03 | 1.44 |
| My worker union |  | 0.99 | 0.92 | 1.06 | 0.89 | 0.78 | 1.02 |
| Were unwell for two or more consecutive days (whether or not worked)? |  | 1.12 | 0.99 | 1.26 | 1.23 | 0.98 | 1.54 |

\* adjusted for age, education, income, children living at home, phase of stay-at-home order, general health; not all effect estimates for support shown; effect estimates for missing categories are not shown
\*\* estimates are per 25 units on Likert-like scale

### Associations with industry and occupation

Industry of employment during the epidemic was associated with anxiety and depression, even after accounting for demographics (**Table 5**). We did not examine effects of work-related factors studied in **Tables 1b** and **2** jointly with industry, because we did not have sufficient data to do so even for the most common industries in our sample (healthcare and higher education) even before restricting to those who reported change in employment during the epidemic. Both crude rates and unadjusted effect estimates on HADS scores in negative binomial regressions are presented; note is made where adjustment for demographics made a notable difference in association with HADS scores.

**Table 5:**
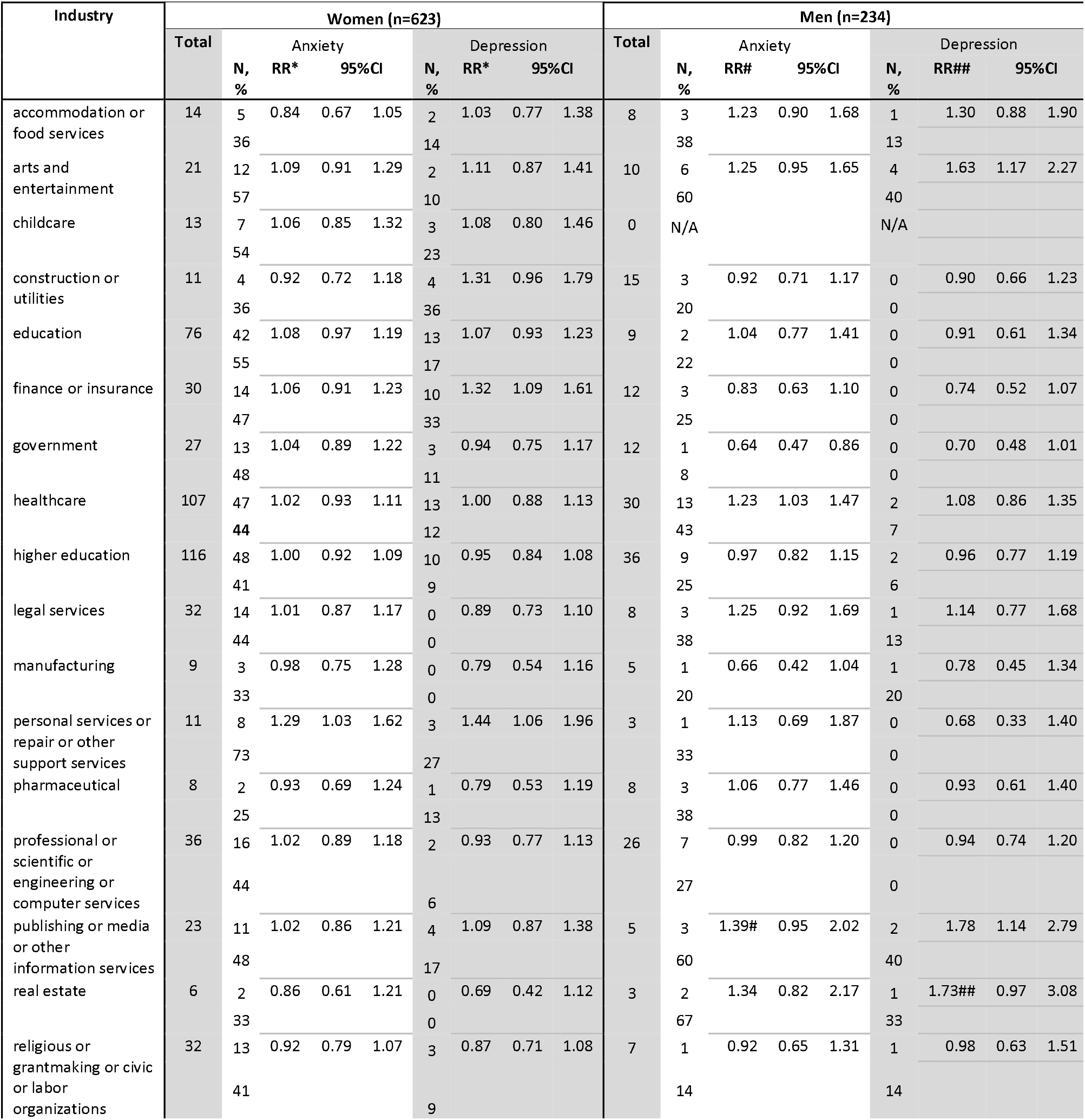
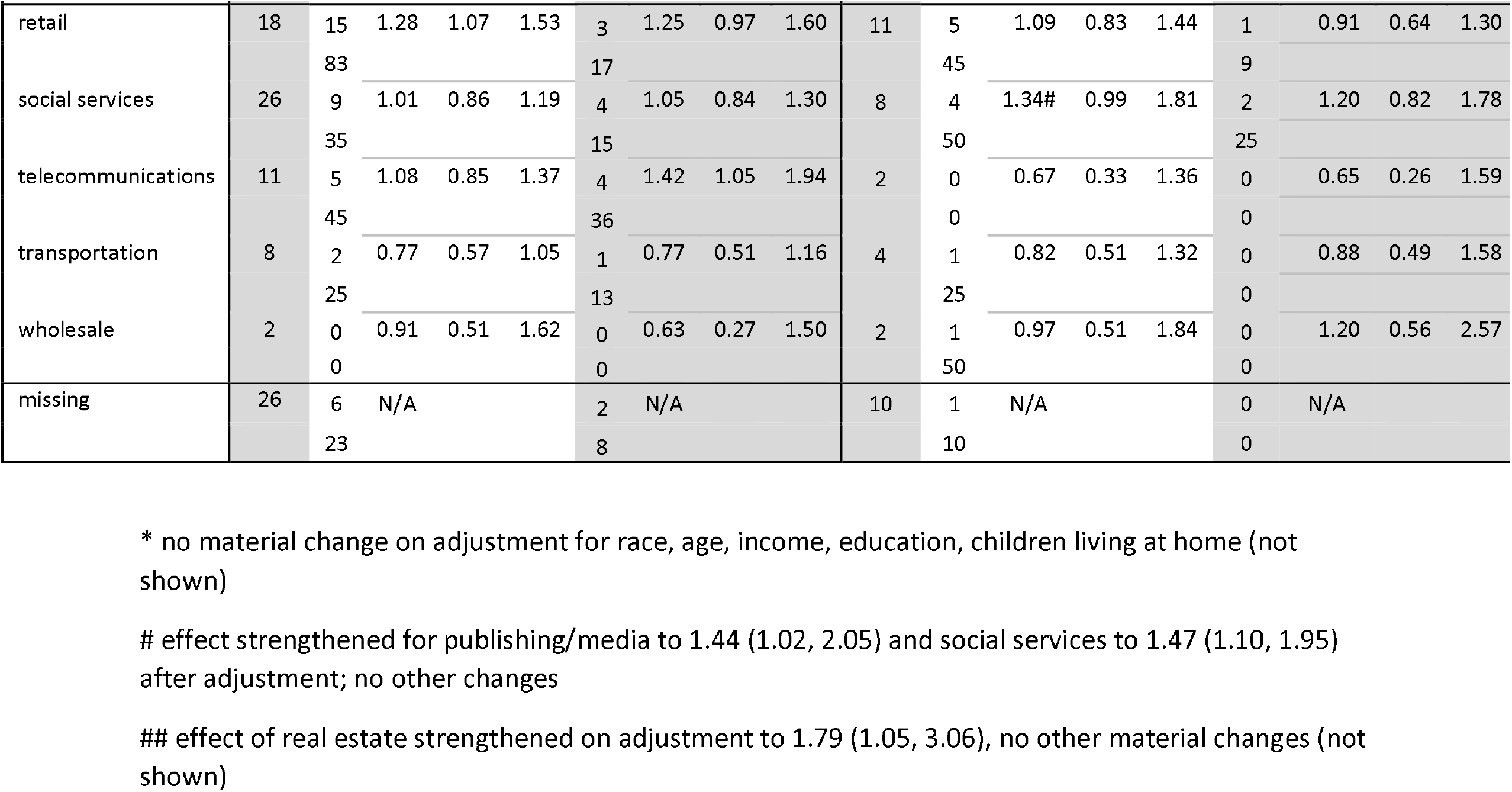
Industry of employment during epidemic and rate of anxiety and depression, along with relative rate (RR) and 95% confidence intervals (CI) of the change in continuous HADS scores relative to sample mean symptoms scores.

Among women, the highest rate of anxiety (83%) was reported within retail industry, based on 15 cases with crude RR 1.28 (95%CI 1.07, 1.53) in comparison to unweighted average across other industries on the continuous HADS anxiety scale (**Table 5**). We observed an association of comparable strength for anxiety among women with work in personal services (8 cases; crude RR 1.29, 95%CI 1.03, 1.62), and a weaker one for education (42 cases; crude RR 1.08, 95%CI 0.97, 1.19). There was a suggestion of deficit in symptoms of anxiety among women working in accommodation or food services (5 cases; RR 0.84, 95%CI 0.67, 1.05) and transportation (2 cases; RR 0.77, 95%CI 0.57, 1.05).

The highest rates of depression (36%) were recorded among women employed in construction and utilities (4 cases; RR 1.31, 95%CI 0.96, 1.79), and telecommunications (4 cases; RR 1.42, 95%CI 1.05, 1.94) (**Table 5**). As with anxiety, there was evidence of excess depression among those employed in retail (3 cases; RR 1.25, 95% 0.97, 1.60) and personal services (3 cases; RR 1.44, 95%CI 1.06, 1.96). The evidence for the association of depression among women with work in insurance and finance was stronger than that for anxiety (10 cases, RR 1.32, 95%CI 1.09, 1.61). There was no robust evidence of reduced risk for other industries, including those with no cases of depression.

Among men, the highest rate of anxiety (67%) was observed in real estate (2 cases; RR 1.34, 95%CI 0.82, 2.17) (**Table 5**). However, the evidence of excess of risk of anxiety is the strongest among men who worked in healthcare, with the rate like that among women in the same sector (43%), based on 13 cases with crude RR 1.22, 95%CI 1.02, 1.46. Men who worked in publishing/media (3 cases; RR 1.39, 95%CI 0.95, 2.02) and social services (4 cases; RR 1.34 (95%CI 0.99, 1.81), were likewise at an increased risk of anxiety; these observations were bolstered by adjustment for demographics. There was a suggestion of lower-than-average risk of anxiety among men who worked in government (1 case; RR 0.64 95%CI 0.47, 0.86) and manufacturing (1 case; RR 0.66, 95%CI 0.42, 1.04).

The highest rates of depression of 40% were among men who worked in arts and entertainment (4 cases; RR 1.63, 95%CI 1.17, 2.27) and publishing/media (2 cases, RR 1.78, 1.14, 2.79). There was also evidence of higher than sample average levels of depression among men who worked in real estate (1 case; crude RR 1.73 95%CI 0.97, 3.08), adjusted RR 1.79 (95% 1.05, 3.06). There were no cases of depression among the 12 men who worked for government and they had lower than average rate of depression symptoms (RR 0.70, 95%CI 0.48, 1.01); this trend is confirmed by examination of continuous HADS scores in **Supplemental Material 2**.

Associations with occupations were weaker than those with industries and can be found in **Supplemental Material 4**. They do suggest elevated anxiety and depression among artists and performers, heightened rates of anxiety among female managers only, and excess of anxiety and depression among women in retails and sales jobs. We noted deficits of (a) depression among women but not men in both science occupations and lawyers/legal occupations, and (b) anxiety among male teachers. These observations were not affected by adjustment for demographics that appeared to play a role in **Table 1a**.

Detailed description of continuous HADS scores by industry and occupation are in **Supplemental Material 2**; they agree with the results summarized above.

## Discussion

We observed variation in prevalence of symptoms of anxiety and depression across jobs and work-related factors in a sample of Philadelphia residents during the first wave of COVID-19 epidemic. The most affected persons worked in sectors both directly impacted by the infections and risk of contagion (e.g., healthcare) and those affected by stay-at-home orders that closed businesses (e.g., arts and entertainment, retail, personal services, real estate), and the least affected worked in sectors less disrupted by stay-at-home orders (e.g., government, manufacturing; salaried employees with access to sick leave and/or those who report they can rely on trade unions for support). Our observation of increased anxiety and depression among younger persons with lower pre-epidemic income is consistent with this. It is paramount to stress that the levels of anxiety and depression were not materially different between persons employed in settings where they thought they came into direct contact with persons infected with virus that causes COVID-19, except when person tested positive for the virus and/or reported to have had no protection from PPE. Curiously, women and men who worked in healthcare were equally anxious (44-45% rate of anxiety cases), despite expected excess of anxiety in women in general, suggesting occupational causes at least among men. A common thread in work-related correlates of anxiety and depression were loss (especially when there was a delay in obtaining unemployment benefits) or reduction in work since start of epidemic, increased anxiety due to telecommuting, and concerns about return to work. Some gender-specific differences related to less secure employment (hourly), lack of access to sick leave though work, and difficulty balancing work and childcare, with men more affected; there were some gender differences by industry and occupation. Only a minority of people are expected to knowingly encounter person ill with COVID-19 though work (population prevalence 1-3% maximum in the region during the survey;(17) 14/911=1.5% in our sample), with the tangible threat of infection at work reported by about 7% in our sample. Consequently, it is believable that, as in our sample, the changes to work due to efforts to contain the pandemic are the dominant occupation factors in anxiety and depression experienced by residents of Philadelphia.

The key mitigating factors were believing to be able to lean on immediate family and trade unions for support, access to sick leave and unemployment benefits, and use of personal prospective equipment when there is perceived threat of contagion. Women who had one-on-one contact with people at work were less depressed. Curiously, not all reported sources of support were related to reduced risk after accounting for other factors: those who sought support mostly from city government were more anxious and depressed.

Rates of anxiety and depression seen in our survey are far above normative values established in the UK,(18) with median normative scores for anxiety in 5-6 range and for depression about 3. This can be interpreted as combination of pandemic-associated stressors and selection of persons with worse health into our survey. However, the finding is congruent with report by Czeisler et al.(1) of decline in mental health during spring and summer of 2020 for the US as the whole, with the rates of anxiety and depression among the self-identified “essential” and healthcare workers comparable to our findings (about 30-40%). Survey of Burstyn & Holt(19) of healthcare workers in one healthcare system that also employed HADS and was delivered online over similar timeframe in Philadelphia, reported rates of anxiety of 34% in nurses and 19% in physicians, and depression rates of 12% and 5% in nurses and physicians, respectively. These rates are lower than in healthcare setting in our sample for both anxiety (44-45%) but like that for depression (7-12%). This can be interpreted as those in health care participating in our survey being more anxious and implies that similar selection bias exists for other occupations. This was expected as it is natural for persons adversely affected by the pandemic to participate in research more willingly. However, it may also reflect differences in working conditions across healthcare systems in the city and the fact that a some healthcare workers in Burstyn & Holt(19) may have worked and lived outside of Philadelphia (though still in the same region). Arguing against strong selection bias is the fact that most participants reported to have been in good to excellent health, although we cannot discount the possibility that those in poor general health were affected by mood disorders that impacted their working lives and were also more likely to participate (there is an association of self-rated health with anxiety and depression in our data). Our work is like that of Smith et al.(4) in Canada, who investigated a convenience sample of non-healthcare workers and noted rates of anxiety and depression, also in the rage of 30-40%, with consistent findings with respect to loss of employment and fear of infection. Our two samples differ in that ours had far fewer unionized participants due to differences in sampling schemes. Although Smith et al.,(20) using the same methods as for general working population, reported higher rates among healthcare workers in Canada, the difference was not stark, just as in our work. All published work in this realm suffers from biased sampling schemes, and yet internally consistent themes emerge.

The most glaring limitation of our survey is that does not represent all working people in Philadelphia and thus any conclusions must be drawn with the understanding that no matter how internally valid, inferences regarding those not represented in the sample (non-white, with less than college education, with low income) is tenuous. Nonetheless, we offer some observations that may not be modified by key demographics, as they relate to universal fears of contagion and economic insecurities, as well as support from immediate family being beneficial. Cross-sectional design and lack of questions on history of mental health limits ability to draw causal inferences. We did control for general health (though not mental health specifically, as was desirable) and health during epidemic, with reports of having been unwell for two or more days associated with anxiety and depression, as in Burstyn & Holt.(19) We also inquired about income pre-epidemic, changes in work during epidemic, and epidemic-specific events (contact with infected, onset of telecommuting, aggravation of challenges of childcare, PPE) and accounted for them in analysis. Nonetheless, it is impossible to rule out residual confounding and reverse causation (e.g., people with finding themselves in less stable and desirable employment situations following onset of mental health problem that either persists or is aggravated by the epidemic). Although we present results by gender, we did not formally test effect modification and we were not able to account for all work-related factors in a single regression model, both due to sparsity of data in some strata. This approach preserved descriptive nature of the study but limits attribution of effects we observe to specific causes and their interaction with gender. All our data is self-reported and thus is vulnerable to recall bias and correlation in errors between exposure and outcomes.

Despite noted shortcomings of our research, we conclude that there is evidence that the disruption to working lives of residents of Philadelphia by the COVID-19 pandemic is related to risk of anxiety and depression, above and beyond effects of encounters with infected individuals at work. While most of the attention has been focused on burdens borne by essential workers (primarily in healthcare), most persons at elevated risk of anxiety and depression are those whose work was not deemed “essential or life-sustaining” by the state. We are hopeful that our investigation will help minimize harm to mental health of all working people during the pandemic and similar future events. We do not believe it is our place to speculate on how mood disorders are best addressed, whether though provision of mental health services, empowerment of families to support each other, or economic policies.

## Data Availability

Data is protected under Drexel University IRB rules that granted approval to the project.

## Acknowledgements

The authors are deeply indebted to all the participants who responded to survey while learning to live under extreme pressures precipitated by the pandemic. Dr Nicola M Cherry of the University of Alberta generously shared ideas and materials on related research. We wish to thank Todd Wolfson and Briar Smith for their feedback on the survey instrument, Mariela Morales for her assistance with online advertisement and Spanish translation, Guangzi Song and Xi Wang for help with the Chinese translation.

