## Supplemental Material 2 for "Symptoms of anxiety and depression in relation to work patterns during the first wave of the COVID-19 epidemic in Philadelphia PA: a cross-sectional survey"

### All who had a job

#### The MEANS Procedure

##### Gender=Female

| Race | N Obs | Variable | Label | N | Mean | Std Dev | 5th Pctl | 50th Pctl | 95th Pctl | Minimum | Maximum |
|---|---|---|---|---|---|---|---|---|---|---|---|
| white | 551 | AscoreR | HADS Anxiety score | 551 | 10.2232305 | 3.9989868 | 4.0000000 | 10.0000000 | 17.0000000 | 0 | 21.0000000 |
| | | DscoreR | HADS Depression score | 551 | 6.5154265 | 3.6535459 | 1.0000000 | 6.0000000 | 13.0000000 | 0 | 20.0000000 |
| black | 63 | AscoreR | HADS Anxiety score | 63 | 9.2380952 | 4.9667095 | 0 | 9.0000000 | 19.0000000 | 0 | 20.0000000 |
| | | DscoreR | HADS Depression score | 63 | 6.0000000 | 3.8100038 | 1.0000000 | 5.0000000 | 14.0000000 | 0 | 18.0000000 |
| other race | 49 | AscoreR | HADS Anxiety score | 49 | 9.3265306 | 4.3223631 | 2.0000000 | 9.0000000 | 16.0000000 | 0 | 20.0000000 |
| | | DscoreR | HADS Depression score | 49 | 6.7346939 | 3.7736781 | 1.0000000 | 7.0000000 | 14.0000000 | 0 | 15.0000000 |

##### Gender=Male

| Race | N Obs | Variable | Label | N | Mean | Std Dev | 5th Pctl | 50th Pctl | 95th Pctl | Minimum | Maximum |
|---|---|---|---|---|---|---|---|---|---|---|---|
| white | 205 | AscoreR | HADS Anxiety score | 205 | 8.6243902 | 4.1243407 | 2.0000000 | 8.0000000 | 16.0000000 | 0 | 20.0000000 |
| | | DscoreR | HADS Depression score | 205 | 5.5463415 | 3.5012671 | 1.0000000 | 5.0000000 | 12.0000000 | 0 | 18.0000000 |
| black | 10 | AscoreR | HADS Anxiety score | 10 | 6.0000000 | 5.1854497 | 1.0000000 | 4.0000000 | 16.0000000 | 1.0000000 | 16.0000000 |
| | | DscoreR | HADS Depression score | 10 | 3.7000000 | 3.5605867 | 0 | 2.5000000 | 9.0000000 | 0 | 9.0000000 |
| other race | 19 | AscoreR | HADS Anxiety score | 19 | 8.0526316 | 3.8510271 | 1.0000000 | 8.0000000 | 17.0000000 | 1.0000000 | 17.0000000 |
| | | DscoreR | HADS Depression score | 19 | 5.3157895 | 3.1278788 | 0 | 5.0000000 | 13.0000000 | 0 | 13.0000000 |

#### The MEANS Procedure

##### Gender=Female

| Race | N Obs | Variable | Label | N | Mean | Std Dev | 5th Pctl | 50th Pctl | 95th Pctl | Minimum | Maximum |
|---|---|---|---|---|---|---|---|---|---|---|---|
| white | 551 | AscoreR | HADS Anxiety score | 551 | 10.2232305 | 3.9989868 | 4.0000000 | 10.0000000 | 17.0000000 | 0 | 21.0000000 |
| | | DscoreR | HADS Depression score | 551 | 6.5154265 | 3.6535459 | 1.0000000 | 6.0000000 | 13.0000000 | 0 | 20.0000000 |
| black | 63 | AscoreR | HADS Anxiety score | 63 | 9.2380952 | 4.9667095 | 0 | 9.0000000 | 19.0000000 | 0 | 20.0000000 |
| | | DscoreR | HADS Depression score | 63 | 6.0000000 | 3.8100038 | 1.0000000 | 5.0000000 | 14.0000000 | 0 | 18.0000000 |
| other race | 49 | AscoreR | HADS Anxiety score | 49 | 9.3265306 | 4.3223631 | 2.0000000 | 9.0000000 | 16.0000000 | 0 | 20.0000000 |
| | | DscoreR | HADS Depression score | 49 | 6.7346939 | 3.7736781 | 1.0000000 | 7.0000000 | 14.0000000 | 0 | 15.0000000 |

##### Gender=Male

| Race | N Obs | Variable | Label | N | Mean | Std Dev | 5th Pctl | 50th Pctl | 95th Pctl | Minimum | Maximum |
|---|---|---|---|---|---|---|---|---|---|---|---|
| white | 205 | AscoreR | HADS Anxiety score | 205 | 8.6243902 | 4.1243407 | 2.0000000 | 8.0000000 | 16.0000000 | 0 | 20.0000000 |
| | | DscoreR | HADS Depression score | 205 | 5.5463415 | 3.5012671 | 1.0000000 | 5.0000000 | 12.0000000 | 0 | 18.0000000 |
| black | 10 | AscoreR | HADS Anxiety score | 10 | 6.0000000 | 5.1854497 | 1.0000000 | 4.0000000 | 16.0000000 | 1.0000000 | 16.0000000 |
| | | DscoreR | HADS Depression score | 10 | 3.7000000 | 3.5605867 | 0 | 2.5000000 | 9.0000000 | 0 | 9.0000000 |
| other race | 19 | AscoreR | HADS Anxiety score | 19 | 8.0526316 | 3.8510271 | 1.0000000 | 8.0000000 | 17.0000000 | 1.0000000 | 17.0000000 |
| | | DscoreR | HADS Depression score | 19 | 5.3157895 | 3.1278788 | 0 | 5.0000000 | 13.0000000 | 0 | 13.0000000 |

#### The MEANS Procedure

##### Gender=Female

| Age | N Obs | Variable | Label | N | Mean | Std Dev | 5th Pctl | 50th Pctl | 95th Pctl | Minimum | Maximum |
|---|---|---|---|---|---|---|---|---|---|---|---|
| less than 35 | 174 | AscoreR<br>DscoreR | HADS Anxiety score<br>HADS Depression score | 174<br>174 | 10.6321839<br>6.5689655 | 4.0619906<br>3.6148957 | 4.0000000<br>1.0000000 | 11.0000000<br>7.0000000 | 18.0000000<br>14.0000000 | 0<br>0 | 19.0000000<br>17.0000000 |
| 35-54 | 292 | AscoreR<br>DscoreR | HADS Anxiety score<br>HADS Depression score | 292<br>292 | 10.0958904<br>6.4315068 | 4.0288116<br>3.4420236 | 4.0000000<br>1.0000000 | 10.0000000<br>6.0000000 | 17.0000000<br>12.0000000 | 0<br>0 | 20.0000000<br>19.0000000 |
| greater than 55 | 197 | AscoreR<br>DscoreR | HADS Anxiety score<br>HADS Depression score | 197<br>197 | 9.5126904<br>6.4822335 | 4.2923866<br>4.0615091 | 2.0000000<br>0 | 9.0000000<br>6.0000000 | 17.0000000<br>14.0000000 | 0<br>0 | 21.0000000<br>20.0000000 |

##### Gender=Male

| Age | N Obs | Variable | Label | N | Mean | Std Dev | 5th Pctl | 50th Pctl | 95th Pctl | Minimum | Maximum |
|---|---|---|---|---|---|---|---|---|---|---|---|
| less than 35 | 36 | AscoreR<br>DscoreR | HADS Anxiety score<br>HADS Depression score | 36<br>36 | 10.3333333<br>6.7777778 | 4.3556203<br>3.0247655 | 3.0000000<br>2.0000000 | 10.0000000<br>6.0000000 | 18.0000000<br>13.0000000 | 1.0000000<br>2.0000000 | 19.0000000<br>13.0000000 |
| 35-54 | 117 | AscoreR<br>DscoreR | HADS Anxiety score<br>HADS Depression score | 117<br>117 | 8.8461538<br>5.6410256 | 4.0292367<br>3.7288751 | 2.0000000<br>1.0000000 | 9.0000000<br>5.0000000 | 16.0000000<br>12.0000000 | 0<br>0 | 20.0000000<br>18.0000000 |
| greater than 55 | 81 | AscoreR<br>DscoreR | HADS Anxiety score<br>HADS Depression score | 81<br>81 | 7.0864198<br>4.5802469 | 3.8800694<br>3.0857098 | 1.0000000<br>1.0000000 | 7.0000000<br>4.0000000 | 14.0000000<br>9.0000000 | 0<br>0 | 17.0000000<br>16.0000000 |

### All who had a job

#### The MEANS Procedure

##### Gender=Female

| Personal income in 2019 | N Obs | Variable | Label | N | Mean | Std Dev | 5th Pctl | 50th Pctl | 95th Pctl | Minimum | Maximum |
|---|---|---|---|---|---|---|---|---|---|---|---|
| less than $40,000 | 132 | AscoreR | HADS Anxiety score | 132 | 10.9469697 | 4.0574395 | 5.0000000 | 11.0000000 | 18.0000000 | 0 | 20.0000000 |
| | | DscoreR | HADS Depression score | 132 | 7.1439394 | 3.5600253 | 2.0000000 | 7.0000000 | 14.0000000 | 0 | 17.0000000 |
| 40,000 - $99,999 | 348 | AscoreR | HADS Anxiety score | 348 | 10.0229885 | 4.1865078 | 4.0000000 | 10.0000000 | 17.0000000 | 0 | 21.0000000 |
| | | DscoreR | HADS Depression score | 348 | 6.2787356 | 3.7354763 | 1.0000000 | 6.0000000 | 13.0000000 | 0 | 20.0000000 |
| $100,000 and above | 172 | AscoreR | HADS Anxiety score | 172 | 9.5988372 | 3.9107886 | 3.0000000 | 9.0000000 | 16.0000000 | 0 | 19.0000000 |
| | | DscoreR | HADS Depression score | 172 | 6.4302326 | 3.6145926 | 1.0000000 | 6.5000000 | 12.0000000 | 0 | 19.0000000 |
| missing | 11 | AscoreR | HADS Anxiety score | 11 | 8.0000000 | 5.1380930 | 0 | 10.0000000 | 14.0000000 | 0 | 14.0000000 |
| | | DscoreR | HADS Depression score | 11 | 5.8181818 | 3.6005050 | 2.0000000 | 5.0000000 | 14.0000000 | 2.0000000 | 14.0000000 |

##### Gender=Male

| Personal income in 2019 | N Obs | Variable | Label | N | Mean | Std Dev | 5th Pctl | 50th Pctl | 95th Pctl | Minimum | Maximum |
|---|---|---|---|---|---|---|---|---|---|---|---|
| less than $40,000 | 34 | AscoreR | HADS Anxiety score | 34 | 9.6470588 | 4.0368181 | 4.0000000 | 9.0000000 | 18.0000000 | 4.0000000 | 20.0000000 |
| | | DscoreR | HADS Depression score | 34 | 6.8235294 | 3.6966044 | 2.0000000 | 6.0000000 | 13.0000000 | 0 | 18.0000000 |
| 40,000 - $99,999 | 96 | AscoreR | HADS Anxiety score | 96 | 9.0520833 | 4.2286486 | 2.0000000 | 9.0000000 | 17.0000000 | 0 | 19.0000000 |
| | | DscoreR | HADS Depression score | 96 | 5.8333333 | 3.6293153 | 1.0000000 | 5.0000000 | 13.0000000 | 0 | 18.0000000 |
| $100,000 and above | 100 | AscoreR | HADS Anxiety score | 100 | 7.4900000 | 4.0163680 | 1.0000000 | 7.0000000 | 14.0000000 | 0 | 18.0000000 |
| | | DscoreR | HADS Depression score | 100 | 4.5700000 | 3.0293347 | 1.0000000 | 4.0000000 | 9.0000000 | 0 | 15.0000000 |
| missing | 4 | AscoreR | HADS Anxiety score | 4 | 8.7500000 | 3.5939764 | 4.0000000 | 9.5000000 | 12.0000000 | 4.0000000 | 12.0000000 |
| | | DscoreR | HADS Depression score | 4 | 6.5000000 | 4.2031734 | 2.0000000 | 6.0000000 | 12.0000000 | 2.0000000 | 12.0000000 |

### The MEANS Procedure

##### Gender=Female

| Education | N Obs | Variable | Label | N | Mean | Std Dev | 5th Pctl | 50th Pctl | 95th Pctl | Minimum | Maximum |
|---|---|---|---|---|---|---|---|---|---|---|---|
| college degree or higher | 589 | AscoreR<br>DscoreR | HADS Anxiety score<br>HADS Depression score | 589<br>589 | 10.0237691<br>6.4108659 | 4.0761600<br>3.5791750 | 4.0000000<br>1.0000000 | 10.0000000<br>6.0000000 | 16.0000000<br>12.0000000 | 0<br>0 | 21.0000000<br>20.0000000 |
| no college degree | 73 | AscoreR<br>DscoreR | HADS Anxiety score<br>HADS Depression score | 73<br>73 | 10.2465753<br>7.0136986 | 4.4621271<br>4.3700151 | 4.0000000<br>0 | 10.0000000<br>6.0000000 | 18.0000000<br>14.0000000 | 0<br>0 | 19.0000000<br>16.0000000 |
| missing | 1 | AscoreR<br>DscoreR | HADS Anxiety score<br>HADS Depression score | 1<br>1 | 20.0000000<br>10.0000000 | .<br>. | 20.0000000<br>10.0000000 | 20.0000000<br>10.0000000 | 20.0000000<br>10.0000000 | 20.0000000<br>10.0000000 | 20.0000000<br>10.0000000 |

##### Gender=Male

| Education | N Obs | Variable | Label | N | Mean | Std Dev | 5th Pctl | 50th Pctl | 95th Pctl | Minimum | Maximum |
|---|---|---|---|---|---|---|---|---|---|---|---|
| college degree or higher | 202 | AscoreR<br>DscoreR | HADS Anxiety score<br>HADS Depression score | 202<br>202 | 8.6237624<br>5.5396040 | 4.1124770<br>3.5226228 | 2.0000000<br>1.0000000 | 8.0000000<br>5.0000000 | 16.0000000<br>12.0000000 | 0<br>0 | 20.0000000<br>18.0000000 |
| no college degree | 29 | AscoreR<br>DscoreR | HADS Anxiety score<br>HADS Depression score | 29<br>29 | 7.6896552<br>5.0689655 | 4.5677710<br>3.2615841 | 0<br>0 | 6.0000000<br>5.0000000 | 17.0000000<br>12.0000000 | 0<br>0 | 17.0000000<br>12.0000000 |
| missing | 3 | AscoreR<br>DscoreR | HADS Anxiety score<br>HADS Depression score | 3<br>3 | 5.3333333<br>3.0000000 | 2.5166115<br>2.0000000 | 3.0000000<br>1.0000000 | 5.0000000<br>3.0000000 | 8.0000000<br>5.0000000 | 3.0000000<br>1.0000000 | 8.0000000<br>5.0000000 |

### All who had a job

#### The MEANS Procedure

##### Gender=Female

| children < 18 years living in your household | N Obs | Variable | Label | N | Mean | Std Dev | 5th Pctl | 50th Pctl | 95th Pctl | Minimum | Maximum |
|---|---|---|---|---|---|---|---|---|---|---|---|
| no | 494 | AscoreR<br>DscoreR | HADS Anxiety score<br>HADS Depression score | 494<br>494 | 10.1315789<br>6.5364372 | 4.1970837<br>3.7651234 | 4.0000000<br>1.0000000 | 10.0000000<br>6.0000000 | 17.0000000<br>13.0000000 | 0<br>0 | 21.0000000<br>20.0000000 |
| yes | 166 | AscoreR<br>DscoreR | HADS Anxiety score<br>HADS Depression score | 166<br>166 | 9.8734940<br>6.3614458 | 3.9591424<br>3.4150409 | 4.0000000<br>1.0000000 | 10.0000000<br>6.0000000 | 18.0000000<br>12.0000000 | 0<br>0 | 20.0000000<br>18.0000000 |
| missing | 3 | AscoreR<br>DscoreR | HADS Anxiety score<br>HADS Depression score | 3<br>3 | 9.3333333<br>4.3333333 | 3.5118846<br>2.5166115 | 6.0000000<br>2.0000000 | 9.0000000<br>4.0000000 | 13.0000000<br>7.0000000 | 6.0000000<br>2.0000000 | 13.0000000<br>7.0000000 |

##### Gender=Male

| children < 18 years living in your household | N Obs | Variable | Label | N | Mean | Std Dev | 5th Pctl | 50th Pctl | 95th Pctl | Minimum | Maximum |
|---|---|---|---|---|---|---|---|---|---|---|---|
| no | 168 | AscoreR<br>DscoreR | HADS Anxiety score<br>HADS Depression score | 168<br>168 | 8.5714286<br>5.4940476 | 4.2434471<br>3.5376453 | 2.0000000<br>1.0000000 | 8.0000000<br>5.0000000 | 17.0000000<br>12.0000000 | 0<br>0 | 20.0000000<br>18.0000000 |
| yes | 62 | AscoreR<br>DscoreR | HADS Anxiety score<br>HADS Depression score | 62<br>62 | 8.3709677<br>5.4354839 | 3.9968589<br>3.3857027 | 2.0000000<br>1.0000000 | 8.5000000<br>5.0000000 | 14.0000000<br>12.0000000 | 0<br>0 | 17.0000000<br>14.0000000 |
| missing | 4 | AscoreR<br>DscoreR | HADS Anxiety score<br>HADS Depression score | 4<br>4 | 5.5000000<br>3.7500000 | 3.1091264<br>2.7537853 | 1.0000000<br>1.0000000 | 6.5000000<br>3.5000000 | 8.0000000<br>7.0000000 | 1.0000000<br>1.0000000 | 8.0000000<br>7.0000000 |

#### The MEANS Procedure

##### Gender=Female

| Marital status | N Obs | Variable | Label | N | Mean | Std Dev | 5th Pctl | 50th Pctl | 95th Pctl | Minimum | Maximum |
|---|---|---|---|---|---|---|---|---|---|---|---|
| married or living as married | 381 | AscoreR<br>DscoreR | HADS Anxiety score<br>HADS Depression score | 381<br>381 | 10.1732283<br>6.5774278 | 3.9571943<br>3.5164897 | 4.0000000<br>1.0000000 | 10.0000000<br>6.0000000 | 16.0000000<br>13.0000000 | 0<br>0 | 21.0000000<br>18.0000000 |
| single | 212 | AscoreR<br>DscoreR | HADS Anxiety score<br>HADS Depression score | 212<br>212 | 9.9811321<br>6.3443396 | 4.3789266<br>3.8820667 | 3.0000000<br>1.0000000 | 10.0000000<br>6.0000000 | 18.0000000<br>14.0000000 | 0<br>0 | 21.0000000<br>20.0000000 |
| widowed, divorced | 67 | AscoreR<br>DscoreR | HADS Anxiety score<br>HADS Depression score | 67<br>67 | 9.6716418<br>6.3134328 | 4.4017492<br>3.9589138 | 3.0000000<br>1.0000000 | 9.0000000<br>6.0000000 | 18.0000000<br>15.0000000 | 0<br>0 | 19.0000000<br>18.0000000 |
| missing | 3 | AscoreR<br>DscoreR | HADS Anxiety score<br>HADS Depression score | 3<br>3 | 10.6666667<br>8.0000000 | 2.5166115<br>2.6457513 | 8.0000000<br>5.0000000 | 11.0000000<br>9.0000000 | 13.0000000<br>10.0000000 | 8.0000000<br>5.0000000 | 13.0000000<br>10.0000000 |

##### Gender=Male

| Marital status | N Obs | Variable | Label | N | Mean | Std Dev | 5th Pctl | 50th Pctl | 95th Pctl | Minimum | Maximum |
|---|---|---|---|---|---|---|---|---|---|---|---|
| married or living as married | 152 | AscoreR<br>DscoreR | HADS Anxiety score<br>HADS Depression score | 152<br>152 | 8.2697368<br>5.2500000 | 4.0214168<br>3.4817404 | 1.0000000<br>1.0000000 | 8.0000000<br>5.0000000 | 16.0000000<br>12.0000000 | 0<br>0 | 18.0000000<br>18.0000000 |
| single | 64 | AscoreR<br>DscoreR | HADS Anxiety score<br>HADS Depression score | 64<br>64 | 9.0468750<br>5.9531250 | 4.4665589<br>3.5296013 | 2.0000000<br>1.0000000 | 8.5000000<br>6.0000000 | 17.0000000<br>12.0000000 | 0<br>0 | 19.0000000<br>18.0000000 |
| widowed, divorced | 15 | AscoreR<br>DscoreR | HADS Anxiety score<br>HADS Depression score | 15<br>15 | 7.4000000<br>5.2666667 | 3.4392690<br>3.3051187 | 4.0000000<br>0 | 6.0000000<br>4.0000000 | 15.0000000<br>12.0000000 | 4.0000000<br>0 | 15.0000000<br>12.0000000 |
| missing | 3 | AscoreR<br>DscoreR | HADS Anxiety score<br>HADS Depression score | 3<br>3 | 11.3333333<br>5.6666667 | 7.5718778<br>4.0414519 | 6.0000000<br>2.0000000 | 8.0000000<br>5.0000000 | 20.0000000<br>10.0000000 | 6.0000000<br>2.0000000 | 20.0000000<br>10.0000000 |

#### The MEANS Procedure

**Gender=Female**

| Employement status at time of survey | N Obs | Variable | Label | N | Mean | Std Dev | 5th Pctl | 50th Pctl | 95th Pctl | Minimum | Maximum |
|---|---|---|---|---|---|---|---|---|---|---|---|
| salaried | 433 | AscoreR<br>DscoreR | HADS Anxiety score<br>HADS Depression score | 433<br>433 | 10.0877598<br>6.3256351 | 4.0821044<br>3.5943260 | 4.0000000<br>1.0000000 | 10.0000000<br>6.0000000 | 17.0000000<br>13.0000000 | 0<br>0 | 21.0000000<br>20.0000000 |
| hourly | 124 | AscoreR<br>DscoreR | HADS Anxiety score<br>HADS Depression score | 124<br>124 | 10.0080645<br>6.8951613 | 4.3101242<br>4.0561477 | 3.0000000<br>1.0000000 | 10.0000000<br>7.0000000 | 18.0000000<br>14.0000000 | 0<br>0 | 19.0000000<br>18.0000000 |
| contractor | 67 | AscoreR<br>DscoreR | HADS Anxiety score<br>HADS Depression score | 67<br>67 | 10.2835821<br>6.9701493 | 4.2455180<br>3.3027544 | 4.0000000<br>3.0000000 | 9.0000000<br>6.0000000 | 19.0000000<br>14.0000000 | 3.0000000<br>1.0000000 | 20.0000000<br>16.0000000 |
| other | 34 | AscoreR<br>DscoreR | HADS Anxiety score<br>HADS Depression score | 34<br>34 | 9.4411765<br>5.7352941 | 4.1792502<br>3.7197982 | 3.0000000<br>0 | 9.0000000<br>6.0000000 | 18.0000000<br>14.0000000 | 2.0000000<br>0 | 21.0000000<br>14.0000000 |
| missing | 5 | AscoreR<br>DscoreR | HADS Anxiety score<br>HADS Depression score | 5<br>5 | 10.6000000<br>8.4000000 | 3.0495901<br>4.3931765 | 7.0000000<br>4.0000000 | 11.0000000<br>7.0000000 | 14.0000000<br>14.0000000 | 7.0000000<br>4.0000000 | 14.0000000<br>14.0000000 |

**Gender=Male**

| Employement status at time of survey | N Obs | Variable | Label | N | Mean | Std Dev | 5th Pctl | 50th Pctl | 95th Pctl | Minimum | Maximum |
|---|---|---|---|---|---|---|---|---|---|---|---|
| salaried | 155 | AscoreR<br>DscoreR | HADS Anxiety score<br>HADS Depression score | 155<br>155 | 8.3870968<br>5.1419355 | 3.9843641<br>3.2402475 | 2.0000000<br>1.0000000 | 8.0000000<br>5.0000000 | 15.0000000<br>12.0000000 | 0<br>0 | 19.0000000<br>18.0000000 |
| hourly | 36 | AscoreR<br>DscoreR | HADS Anxiety score<br>HADS Depression score | 36<br>36 | 9.0000000<br>5.7777778 | 4.9338481<br>3.4151855 | 1.0000000<br>0 | 8.0000000<br>6.0000000 | 17.0000000<br>12.0000000 | 0<br>0 | 20.0000000<br>12.0000000 |
| contractor | 34 | AscoreR<br>DscoreR | HADS Anxiety score<br>HADS Depression score | 34<br>34 | 8.1176471<br>6.3529412 | 3.8906625<br>4.3126477 | 4.0000000<br>0 | 7.0000000<br>5.0000000 | 16.0000000<br>16.0000000 | 2.0000000<br>0 | 17.0000000<br>18.0000000 |
| other | 9 | AscoreR<br>DscoreR | HADS Anxiety score<br>HADS Depression score | 9<br>9 | 9.0000000<br>6.0000000 | 5.4083269<br>4.1231056 | 1.0000000<br>1.0000000 | 10.0000000<br>6.0000000 | 18.0000000<br>15.0000000 | 1.0000000<br>1.0000000 | 18.0000000<br>15.0000000 |

### The MEANS Procedure

#### Gender=Female

| worked 5+ days a week | N Obs | Variable | Label | N | Mean | Std Dev | 5th Pctl | 50th Pctl | 95th Pctl | Minimum | Maximum |
|---|---|---|---|---|---|---|---|---|---|---|---|
| <5 days a week | 166 | AscoreR<br>DscoreR | HADS Anxiety score<br>HADS Depression score | 166<br>166 | 9.6987952<br>6.5481928 | 4.2290050<br>3.8573931 | 3.0000000<br>1.0000000 | 9.0000000<br>6.0000000 | 16.0000000<br>14.0000000 | 0<br>0 | 21.0000000<br>20.0000000 |
| 5+ days a week | 497 | AscoreR<br>DscoreR | HADS Anxiety score<br>HADS Depression score | 497<br>497 | 10.1851107<br>6.4607646 | 4.0968541<br>3.6165739 | 4.0000000<br>1.0000000 | 10.0000000<br>6.0000000 | 18.0000000<br>13.0000000 | 0<br>0 | 21.0000000<br>19.0000000 |

#### Gender=Male

| worked 5+ days a week | N Obs | Variable | Label | N | Mean | Std Dev | 5th Pctl | 50th Pctl | 95th Pctl | Minimum | Maximum |
|---|---|---|---|---|---|---|---|---|---|---|---|
| <5 days a week | 44 | AscoreR<br>DscoreR | HADS Anxiety score<br>HADS Depression score | 44<br>44 | 7.7954545<br>5.5227273 | 3.6317690<br>3.5795845 | 2.0000000<br>1.0000000 | 8.0000000<br>5.0000000 | 13.0000000<br>12.0000000 | 0<br>1.0000000 | 18.0000000<br>17.0000000 |
| 5+ days a week | 190 | AscoreR<br>DscoreR | HADS Anxiety score<br>HADS Depression score | 190<br>190 | 8.6210526<br>5.4315789 | 4.2766227<br>3.4676208 | 2.0000000<br>1.0000000 | 8.5000000<br>5.0000000 | 16.0000000<br>12.0000000 | 0<br>0 | 20.0000000<br>18.0000000 |

#### The MEANS Procedure

##### Gender=Female

| Race | N Obs | Variable | Label | N | Mean | Std Dev | 5th Pctl | 50th Pctl | 95th Pctl | Minimum | Maximum |
|---|---|---|---|---|---|---|---|---|---|---|---|
| white | 551 | AscoreR | HADS Anxiety score | 551 | 10.2232305 | 3.9989868 | 4.0000000 | 10.0000000 | 17.0000000 | 0 | 21.0000000 |
| | | DscoreR | HADS Depression score | 551 | 6.5154265 | 3.6535459 | 1.0000000 | 6.0000000 | 13.0000000 | 0 | 20.0000000 |
| black | 63 | AscoreR | HADS Anxiety score | 63 | 9.2380952 | 4.9667095 | 0 | 9.0000000 | 19.0000000 | 0 | 20.0000000 |
| | | DscoreR | HADS Depression score | 63 | 6.0000000 | 3.8100038 | 1.0000000 | 5.0000000 | 14.0000000 | 0 | 18.0000000 |
| other race | 49 | AscoreR | HADS Anxiety score | 49 | 9.3265306 | 4.3223631 | 2.0000000 | 9.0000000 | 16.0000000 | 0 | 20.0000000 |
| | | DscoreR | HADS Depression score | 49 | 6.7346939 | 3.7736781 | 1.0000000 | 7.0000000 | 14.0000000 | 0 | 15.0000000 |

##### Gender=Male

| Race | N Obs | Variable | Label | N | Mean | Std Dev | 5th Pctl | 50th Pctl | 95th Pctl | Minimum | Maximum |
|---|---|---|---|---|---|---|---|---|---|---|---|
| white | 205 | AscoreR | HADS Anxiety score | 205 | 8.6243902 | 4.1243407 | 2.0000000 | 8.0000000 | 16.0000000 | 0 | 20.0000000 |
| | | DscoreR | HADS Depression score | 205 | 5.5463415 | 3.5012671 | 1.0000000 | 5.0000000 | 12.0000000 | 0 | 18.0000000 |
| black | 10 | AscoreR | HADS Anxiety score | 10 | 6.0000000 | 5.1854497 | 1.0000000 | 4.0000000 | 16.0000000 | 1.0000000 | 16.0000000 |
| | | DscoreR | HADS Depression score | 10 | 3.7000000 | 3.5605867 | 0 | 2.5000000 | 9.0000000 | 0 | 9.0000000 |
| other race | 19 | AscoreR | HADS Anxiety score | 19 | 8.0526316 | 3.8510271 | 1.0000000 | 8.0000000 | 17.0000000 | 1.0000000 | 17.0000000 |
| | | DscoreR | HADS Depression score | 19 | 5.3157895 | 3.1278788 | 0 | 5.0000000 | 13.0000000 | 0 | 13.0000000 |

### The MEANS Procedure

##### Gender=Female

| worked 40+ hours a week | N Obs | Variable | Label | N | Mean | Std Dev | 5th Pctl | 50th Pctl | 95th Pctl | Minimum | Maximum |
|---|---|---|---|---|---|---|---|---|---|---|---|
| <40 hrs a week | 298 | AscoreR<br>DscoreR | HADS Anxiety score<br>HADS Depression score | 298<br>298 | 9.8959732<br>6.3791946 | 4.1502810<br>3.6909896 | 3.0000000<br>1.0000000 | 10.0000000<br>6.0000000 | 18.0000000<br>14.0000000 | 0<br>0 | 21.0000000<br>20.0000000 |
| 40+ hrs a week | 365 | AscoreR<br>DscoreR | HADS Anxiety score<br>HADS Depression score | 365<br>365 | 10.2000000<br>6.5671233 | 4.1185722<br>3.6658528 | 4.0000000<br>1.0000000 | 10.0000000<br>7.0000000 | 17.0000000<br>13.0000000 | 0<br>0 | 21.0000000<br>18.0000000 |

##### Gender=Male

| worked 40+ hours a week | N Obs | Variable | Label | N | Mean | Std Dev | 5th Pctl | 50th Pctl | 95th Pctl | Minimum | Maximum |
|---|---|---|---|---|---|---|---|---|---|---|---|
| <40 hrs a week | 80 | AscoreR<br>DscoreR | HADS Anxiety score<br>HADS Depression score | 80<br>80 | 8.1500000<br>5.4625000 | 4.2039764<br>3.5930444 | 2.0000000<br>1.0000000 | 7.0000000<br>5.0000000 | 16.0000000<br>12.0000000 | 0<br>0 | 19.0000000<br>18.0000000 |
| 40+ hrs a week | 154 | AscoreR<br>DscoreR | HADS Anxiety score<br>HADS Depression score | 154<br>154 | 8.6298701<br>5.4415584 | 4.1538254<br>3.4337559 | 2.0000000<br>1.0000000 | 9.0000000<br>5.0000000 | 16.0000000<br>12.0000000 | 0<br>0 | 20.0000000<br>18.0000000 |

#### The MEANS Procedure

##### Gender=Female

| Occupational one-on-one contact with known or suspected people with Covid-19 | N Obs | Variable | Label | N | Mean | Std Dev | 5th Pctl | 50th Pctl | 95th Pctl | Minimum | Maximum |
|---|---|---|---|---|---|---|---|---|---|---|---|
| no | 513 | AscoreR<br>DscoreR | HADS Anxiety score<br>HADS Depression score | 513<br>513 | 10.0721248<br>6.5360624 | 4.1936362<br>3.7248067 | 4.0000000<br>1.0000000 | 10.0000000<br>6.0000000 | 18.0000000<br>13.0000000 | 0<br>0 | 21.0000000<br>20.0000000 |
| yes | 48 | AscoreR<br>DscoreR | HADS Anxiety score<br>HADS Depression score | 48<br>48 | 9.8958333<br>6.0000000 | 4.7991115<br>3.6727866 | 1.0000000<br>0 | 9.0000000<br>6.0000000 | 19.0000000<br>12.0000000 | 0<br>0 | 20.0000000<br>14.0000000 |
| do not know | 102 | AscoreR<br>DscoreR | HADS Anxiety score<br>HADS Depression score | 102<br>102 | 10.0980392<br>6.4411765 | 3.4712676<br>3.4370536 | 5.0000000<br>2.0000000 | 10.0000000<br>6.0000000 | 16.0000000<br>12.0000000 | 3.0000000<br>0 | 18.0000000<br>17.0000000 |

##### Gender=Male

| Occupational one-on-one contact with known or suspected people with Covid-19 | N Obs | Variable | Label | N | Mean | Std Dev | 5th Pctl | 50th Pctl | 95th Pctl | Minimum | Maximum |
|---|---|---|---|---|---|---|---|---|---|---|---|
| no | 181 | AscoreR<br>DscoreR | HADS Anxiety score<br>HADS Depression score | 181<br>181 | 8.2154696<br>5.4475138 | 3.8228819<br>3.3737149 | 2.0000000<br>1.0000000 | 8.0000000<br>5.0000000 | 14.0000000<br>12.0000000 | 0<br>0 | 19.0000000<br>18.0000000 |
| yes | 20 | AscoreR<br>DscoreR | HADS Anxiety score<br>HADS Depression score | 20<br>20 | 8.8000000<br>5.6000000 | 4.8406176<br>4.0183788 | 1.0000000<br>1.0000000 | 8.0000000<br>5.0000000 | 17.0000000<br>13.5000000 | 1.0000000<br>1.0000000 | 17.0000000<br>17.0000000 |
| do not know | 33 | AscoreR<br>DscoreR | HADS Anxiety score<br>HADS Depression score | 33<br>33 | 9.6363636<br>5.3636364 | 5.3433263<br>3.8227786 | 0<br>0 | 9.0000000<br>5.0000000 | 18.0000000<br>14.0000000 | 0<br>0 | 20.0000000<br>15.0000000 |

### All who had a job

#### The MEANS Procedure

##### Gender=Female

| Occupational one-on-one contact with known or suspected people with Covid-19 | N Obs | Variable | Label | N | Mean | Std Dev | 5th Pctl | 50th Pctl | 95th Pctl | Minimum | Maximum |
|---|---|---|---|---|---|---|---|---|---|---|---|
| no | 513 | AscoreR<br>DscoreR | HADS Anxiety score<br>HADS Depression score | 513<br>513 | 10.0721248<br>6.5360624 | 4.1936362<br>3.7248067 | 4.0000000<br>1.0000000 | 10.0000000<br>6.0000000 | 18.0000000<br>13.0000000 | 0<br>0 | 21.0000000<br>20.0000000 |
| yes | 48 | AscoreR<br>DscoreR | HADS Anxiety score<br>HADS Depression score | 48<br>48 | 9.8958333<br>6.0000000 | 4.7991115<br>3.6727866 | 1.0000000<br>0 | 9.0000000<br>6.0000000 | 19.0000000<br>12.0000000 | 0<br>0 | 20.0000000<br>14.0000000 |
| do not know | 102 | AscoreR<br>DscoreR | HADS Anxiety score<br>HADS Depression score | 102<br>102 | 10.0980392<br>6.4411765 | 3.4712676<br>3.4370536 | 5.0000000<br>2.0000000 | 10.0000000<br>6.0000000 | 16.0000000<br>12.0000000 | 3.0000000<br>0 | 18.0000000<br>17.0000000 |

##### Gender=Male

| Occupational one-on-one contact with known or suspected people with Covid-19 | N Obs | Variable | Label | N | Mean | Std Dev | 5th Pctl | 50th Pctl | 95th Pctl | Minimum | Maximum |
|---|---|---|---|---|---|---|---|---|---|---|---|
| no | 181 | AscoreR<br>DscoreR | HADS Anxiety score<br>HADS Depression score | 181<br>181 | 8.2154696<br>5.4475138 | 3.8228819<br>3.3737149 | 2.0000000<br>1.0000000 | 8.0000000<br>5.0000000 | 14.0000000<br>12.0000000 | 0<br>0 | 19.0000000<br>18.0000000 |
| yes | 20 | AscoreR<br>DscoreR | HADS Anxiety score<br>HADS Depression score | 20<br>20 | 8.8000000<br>5.6000000 | 4.8406176<br>4.0183788 | 1.0000000<br>1.0000000 | 8.0000000<br>5.0000000 | 17.0000000<br>13.5000000 | 1.0000000<br>1.0000000 | 17.0000000<br>17.0000000 |
| do not know | 33 | AscoreR<br>DscoreR | HADS Anxiety score<br>HADS Depression score | 33<br>33 | 9.6363636<br>5.3636364 | 5.3433263<br>3.8227786 | 0<br>0 | 9.0000000<br>5.0000000 | 18.0000000<br>14.0000000 | 0<br>0 | 20.0000000<br>15.0000000 |

#### The MEANS Procedure

##### Gender=Female

| Job designated as essential or life-sustaining | N Obs | Variable | Label | N | Mean | Std Dev | 5th Pctl | 50th Pctl | 95th Pctl | Minimum | Maximum |
|---|---|---|---|---|---|---|---|---|---|---|---|
| no | 485 | AscoreR<br>DscoreR | HADS Anxiety score<br>HADS Depression score | 485<br>485 | 10.0824742<br>6.5237113 | 4.1187690<br>3.6491189 | 4.0000000<br>1.0000000 | 10.0000000<br>6.0000000 | 18.0000000<br>13.0000000 | 0<br>0 | 21.0000000<br>19.0000000 |
| yes | 126 | AscoreR<br>DscoreR | HADS Anxiety score<br>HADS Depression score | 126<br>126 | 9.8412698<br>6.1507937 | 4.1654055<br>3.7225098 | 4.0000000<br>1.0000000 | 9.0000000<br>6.0000000 | 16.0000000<br>12.0000000 | 0<br>0 | 20.0000000<br>18.0000000 |
| maybe | 52 | AscoreR<br>DscoreR | HADS Anxiety score<br>HADS Depression score | 52<br>52 | 10.4230769<br>6.9038462 | 4.2303579<br>3.8156521 | 5.0000000<br>1.0000000 | 10.0000000<br>7.0000000 | 18.0000000<br>13.0000000 | 0<br>0 | 21.0000000<br>20.0000000 |

##### Gender=Male

| Job designated as essential or life-sustaining | N Obs | Variable | Label | N | Mean | Std Dev | 5th Pctl | 50th Pctl | 95th Pctl | Minimum | Maximum |
|---|---|---|---|---|---|---|---|---|---|---|---|
| no | 151 | AscoreR<br>DscoreR | HADS Anxiety score<br>HADS Depression score | 151<br>151 | 8.4039735<br>5.4966887 | 3.8870362<br>3.4464942 | 2.0000000<br>1.0000000 | 8.0000000<br>5.0000000 | 14.0000000<br>12.0000000 | 0<br>0 | 19.0000000<br>18.0000000 |
| yes | 70 | AscoreR<br>DscoreR | HADS Anxiety score<br>HADS Depression score | 70<br>70 | 8.3857143<br>5.1857143 | 4.6758487<br>3.6407232 | 1.0000000<br>0 | 8.0000000<br>5.0000000 | 17.0000000<br>12.0000000 | 0<br>0 | 20.0000000<br>17.0000000 |
| maybe | 13 | AscoreR<br>DscoreR | HADS Anxiety score<br>HADS Depression score | 13<br>13 | 9.6153846<br>6.3076923 | 4.6104675<br>3.0655238 | 1.0000000<br>2.0000000 | 10.0000000<br>6.0000000 | 17.0000000<br>11.0000000 | 1.0000000<br>2.0000000 | 17.0000000<br>11.0000000 |

### All who had a job

#### The MEANS Procedure

##### Gender=Female

| Used PPE at work | N Obs | Variable | Label | N | Mean | Std Dev | 5th Pctl | 50th Pctl | 95th Pctl | Minimum | Maximum |
|---|---|---|---|---|---|---|---|---|---|---|---|
| no | 378 | AscoreR | HADS Anxiety score | 378 | 10.2592593 | 4.1664928 | 4.0000000 | 10.0000000 | 18.0000000 | 0 | 21.0000000 |
| | | DscoreR | HADS Depression score | 378 | 6.7671958 | 3.7648075 | 1.0000000 | 7.0000000 | 14.0000000 | 0 | 20.0000000 |
| yes | 183 | AscoreR | HADS Anxiety score | 183 | 9.6393443 | 4.3829068 | 2.0000000 | 9.0000000 | 17.0000000 | 0 | 20.0000000 |
| | | DscoreR | HADS Depression score | 183 | 5.9180328 | 3.5686136 | 0 | 6.0000000 | 12.0000000 | 0 | 18.0000000 |
| missing | 102 | AscoreR | HADS Anxiety score | 102 | 10.0980392 | 3.4712676 | 5.0000000 | 10.0000000 | 16.0000000 | 3.0000000 | 18.0000000 |
| | | DscoreR | HADS Depression score | 102 | 6.4411765 | 3.4370536 | 2.0000000 | 6.0000000 | 12.0000000 | 0 | 17.0000000 |

##### Gender=Male

| Used PPE at work | N Obs | Variable | Label | N | Mean | Std Dev | 5th Pctl | 50th Pctl | 95th Pctl | Minimum | Maximum |
|---|---|---|---|---|---|---|---|---|---|---|---|
| no | 134 | AscoreR | HADS Anxiety score | 134 | 8.1567164 | 3.6683046 | 2.0000000 | 8.0000000 | 14.0000000 | 1.0000000 | 18.0000000 |
| | | DscoreR | HADS Depression score | 134 | 5.5746269 | 3.4714233 | 1.0000000 | 5.0000000 | 12.0000000 | 0 | 18.0000000 |
| yes | 67 | AscoreR | HADS Anxiety score | 67 | 8.5074627 | 4.4154451 | 1.0000000 | 8.0000000 | 17.0000000 | 0 | 19.0000000 |
| | | DscoreR | HADS Depression score | 67 | 5.2388060 | 3.3669047 | 1.0000000 | 5.0000000 | 12.0000000 | 0 | 14.0000000 |
| missing | 33 | AscoreR | HADS Anxiety score | 33 | 9.6363636 | 5.3433263 | 0 | 9.0000000 | 18.0000000 | 0 | 20.0000000 |
| | | DscoreR | HADS Depression score | 33 | 5.3636364 | 3.8227786 | 0 | 5.0000000 | 14.0000000 | 0 | 15.0000000 |

#### The MEANS Procedure

**Gender=Female**

| Period unable to work due to high risk of spreading of COVID-19 | N Obs | Variable | Label | N | Mean | Std Dev | 5th Pctl | 50th Pctl | 95th Pctl | Minimum | Maximum |
|---|---|---|---|---|---|---|---|---|---|---|---|
| no | 587 | AscoreR | HADS Anxiety score | 587 | 10.0477002 | 4.0970864 | 4.0000000 | 10.0000000 | 18.0000000 | 0 | 21.0000000 |
| | | DscoreR | HADS Depression score | 587 | 6.4190801 | 3.6992081 | 1.0000000 | 6.0000000 | 13.0000000 | 0 | 20.0000000 |
| yes | 76 | AscoreR | HADS Anxiety score | 76 | 10.1842105 | 4.4233035 | 1.0000000 | 10.0000000 | 16.0000000 | 0 | 20.0000000 |
| | | DscoreR | HADS Depression score | 76 | 6.9736842 | 3.4716900 | 1.0000000 | 7.0000000 | 14.0000000 | 0 | 15.0000000 |

**Gender=Male**

| Period unable to work due to high risk of spreading of COVID-19 | N Obs | Variable | Label | N | Mean | Std Dev | 5th Pctl | 50th Pctl | 95th Pctl | Minimum | Maximum |
|---|---|---|---|---|---|---|---|---|---|---|---|
| no | 212 | AscoreR | HADS Anxiety score | 212 | 8.4575472 | 4.1171344 | 2.0000000 | 8.0000000 | 16.0000000 | 0 | 20.0000000 |
| | | DscoreR | HADS Depression score | 212 | 5.3584906 | 3.3717215 | 1.0000000 | 5.0000000 | 12.0000000 | 0 | 18.0000000 |
| yes | 22 | AscoreR | HADS Anxiety score | 22 | 8.5454545 | 4.7381659 | 1.0000000 | 8.0000000 | 16.0000000 | 1.0000000 | 17.0000000 |
| | | DscoreR | HADS Depression score | 22 | 6.3181818 | 4.4011510 | 2.0000000 | 5.5000000 | 16.0000000 | 1.0000000 | 18.0000000 |

#### The MEANS Procedure

**Gender=Female**

| Period unable to work due to other reasons | N Obs | Variable | Label | N | Mean | Std Dev | 5th Pctl | 50th Pctl | 95th Pctl | Minimum | Maximum |
|---|---|---|---|---|---|---|---|---|---|---|---|
| no | 527 | AscoreR | HADS Anxiety score | 527 | 10.1214421 | 4.2274289 | 4.0000000 | 10.0000000 | 18.0000000 | 0 | 21.0000000 |
| | | DscoreR | HADS Depression score | 527 | 6.5540797 | 3.7356083 | 1.0000000 | 6.0000000 | 13.0000000 | 0 | 20.0000000 |
| yes | 136 | AscoreR | HADS Anxiety score | 136 | 9.8382353 | 3.7480278 | 4.0000000 | 9.5000000 | 16.0000000 | 0 | 19.0000000 |
| | | DscoreR | HADS Depression score | 136 | 6.2058824 | 3.4321306 | 1.0000000 | 6.0000000 | 12.0000000 | 0 | 18.0000000 |

**Gender=Male**

| Period unable to work due to other reasons | N Obs | Variable | Label | N | Mean | Std Dev | 5th Pctl | 50th Pctl | 95th Pctl | Minimum | Maximum |
|---|---|---|---|---|---|---|---|---|---|---|---|
| no | 193 | AscoreR | HADS Anxiety score | 193 | 8.3989637 | 4.1447003 | 2.0000000 | 8.0000000 | 16.0000000 | 0 | 20.0000000 |
| | | DscoreR | HADS Depression score | 193 | 5.4663212 | 3.5178348 | 1.0000000 | 5.0000000 | 12.0000000 | 0 | 18.0000000 |
| yes | 40 | AscoreR | HADS Anxiety score | 40 | 8.8500000 | 4.3474130 | 2.0000000 | 9.0000000 | 17.5000000 | 0 | 18.0000000 |
| | | DscoreR | HADS Depression score | 40 | 5.3000000 | 3.3604029 | 1.0000000 | 5.0000000 | 11.0000000 | 1.0000000 | 18.0000000 |
| 3 | 1 | AscoreR | HADS Anxiety score | 1 | 6.0000000 | . | 6.0000000 | 6.0000000 | 6.0000000 | 6.0000000 | 6.0000000 |
| | | DscoreR | HADS Depression score | 1 | 8.0000000 | . | 8.0000000 | 8.0000000 | 8.0000000 | 8.0000000 | 8.0000000 |

#### The MEANS Procedure

##### Gender=Female

| access to paid sick leave or disability insurance through work | N Obs | Variable | Label | N | Mean | Std Dev | 5th Pctl | 50th Pctl | 95th Pctl | Minimum | Maximum |
|---|---|---|---|---|---|---|---|---|---|---|---|
| no | 133 | AscoreR | HADS Anxiety score | 133 | 9.7142857 | 4.2561389 | 3.0000000 | 9.0000000 | 17.0000000 | 0 | 20.0000000 |
| | | DscoreR | HADS Depression score | 133 | 6.7443609 | 3.5879726 | 1.0000000 | 6.0000000 | 14.0000000 | 0 | 16.0000000 |
| yes | 510 | AscoreR | HADS Anxiety score | 510 | 10.1274510 | 4.0227046 | 4.0000000 | 10.0000000 | 17.0000000 | 0 | 21.0000000 |
| | | DscoreR | HADS Depression score | 510 | 6.3901961 | 3.6891363 | 1.0000000 | 6.0000000 | 13.0000000 | 0 | 20.0000000 |
| unsure | 20 | AscoreR | HADS Anxiety score | 20 | 10.7500000 | 5.8478516 | 0 | 11.0000000 | 19.5000000 | 0 | 20.0000000 |
| | | DscoreR | HADS Depression score | 20 | 7.1000000 | 3.9456839 | 1.0000000 | 7.0000000 | 13.0000000 | 1.0000000 | 13.0000000 |

##### Gender=Male

| access to paid sick leave or disability insurance through work | N Obs | Variable | Label | N | Mean | Std Dev | 5th Pctl | 50th Pctl | 95th Pctl | Minimum | Maximum |
|---|---|---|---|---|---|---|---|---|---|---|---|
| no | 57 | AscoreR | HADS Anxiety score | 57 | 8.3508772 | 4.0773382 | 2.0000000 | 7.0000000 | 16.0000000 | 1.0000000 | 18.0000000 |
| | | DscoreR | HADS Depression score | 57 | 6.5614035 | 4.4642130 | 1.0000000 | 6.0000000 | 17.0000000 | 0 | 18.0000000 |
| yes | 172 | AscoreR | HADS Anxiety score | 172 | 8.4709302 | 4.2197355 | 2.0000000 | 8.0000000 | 17.0000000 | 0 | 20.0000000 |
| | | DscoreR | HADS Depression score | 172 | 5.0813953 | 3.0386020 | 1.0000000 | 5.0000000 | 11.0000000 | 0 | 14.0000000 |
| unsure | 5 | AscoreR | HADS Anxiety score | 5 | 9.6000000 | 4.0373258 | 5.0000000 | 9.0000000 | 16.0000000 | 5.0000000 | 16.0000000 |
| | | DscoreR | HADS Depression score | 5 | 5.4000000 | 2.9664794 | 2.0000000 | 4.0000000 | 9.0000000 | 2.0000000 | 9.0000000 |

#### The MEANS Procedure

**Gender=Female**

| Occupation | N Obs | Variable | Label | N | Mean | Std Dev | 5th Pctl | 50th Pctl | 95th Pctl | Minimum | Maximum |
|---|---|---|---|---|---|---|---|---|---|---|---|
| accountants, financial specialists, and other business operatio | 33 | AscoreR<br>DscoreR | HADS Anxiety score<br>HADS Depression score | 33<br>33 | 9.2727273<br>6.2424242 | 3.7605911<br>2.8942864 | 3.0000000<br>2.0000000 | 8.0000000<br>7.0000000 | 15.0000000<br>11.0000000 | 1.0000000<br>2.0000000 | 18.0000000<br>14.0000000 |
| adminstrative support | 81 | AscoreR<br>DscoreR | HADS Anxiety score<br>HADS Depression score | 81<br>81 | 9.4938272<br>6.0617284 | 4.3391343<br>4.0010801 | 3.0000000<br>1.0000000 | 9.0000000<br>5.0000000 | 17.0000000<br>13.0000000 | 1.0000000<br>0 | 21.0000000<br>20.0000000 |
| architects and engineers | 10 | AscoreR<br>DscoreR | HADS Anxiety score<br>HADS Depression score | 10<br>10 | 9.3000000<br>7.4000000 | 3.8600518<br>3.0983867 | 4.0000000<br>3.0000000 | 8.5000000<br>7.5000000 | 15.0000000<br>12.0000000 | 4.0000000<br>3.0000000 | 15.0000000<br>12.0000000 |
| artists and performers | 19 | AscoreR<br>DscoreR | HADS Anxiety score<br>HADS Depression score | 19<br>19 | 11.1052632<br>8.0000000 | 4.0400046<br>3.8873013 | 5.0000000<br>1.0000000 | 11.0000000<br>8.0000000 | 20.0000000<br>16.0000000 | 5.0000000<br>1.0000000 | 20.0000000<br>16.0000000 |
| computer occupations | 17 | AscoreR<br>DscoreR | HADS Anxiety score<br>HADS Depression score | 17<br>17 | 10.4705882<br>5.8235294 | 4.7581200<br>3.8767785 | 0<br>0 | 11.0000000<br>7.0000000 | 18.0000000<br>13.0000000 | 0<br>0 | 18.0000000<br>13.0000000 |
| counselors, social workers, and community service occupations | 32 | AscoreR<br>DscoreR | HADS Anxiety score<br>HADS Depression score | 32<br>32 | 9.8437500<br>6.2812500 | 3.8697285<br>3.1339388 | 4.0000000<br>2.0000000 | 9.0000000<br>6.0000000 | 15.0000000<br>11.0000000 | 1.0000000<br>0 | 19.0000000<br>14.0000000 |
| healthcare practitioners and technicians | 55 | AscoreR<br>DscoreR | HADS Anxiety score<br>HADS Depression score | 55<br>55 | 9.6363636<br>6.4727273 | 4.3306130<br>3.8483019 | 3.0000000<br>1.0000000 | 9.0000000<br>6.0000000 | 16.0000000<br>12.0000000 | 0<br>0 | 20.0000000<br>17.0000000 |
| laywers and other legal occupations | 28 | AscoreR<br>DscoreR | HADS Anxiety score<br>HADS Depression score | 28<br>28 | 9.3928571<br>5.1785714 | 3.9472380<br>2.9445942 | 3.0000000<br>1.0000000 | 9.0000000<br>5.0000000 | 16.0000000<br>10.0000000 | 1.0000000<br>0 | 18.0000000<br>10.0000000 |
| librarians and curators | 16 | AscoreR<br>DscoreR | HADS Anxiety score<br>HADS Depression score | 16<br>16 | 10.3750000<br>7.0000000 | 3.7036019<br>3.6514837 | 5.0000000<br>0 | 10.0000000<br>6.5000000 | 17.0000000<br>14.0000000 | 5.0000000<br>0 | 17.0000000<br>14.0000000 |

#### The MEANS Procedure

**Gender=Female**

| Occupation | N Obs | Variable | Label | N | Mean | Std Dev | 5th Pctl | 50th Pctl | 95th Pctl | Minimum | Maximum |
|---|---|---|---|---|---|---|---|---|---|---|---|
| life, physical, and social scientists | 57 | AscoreR<br>DscoreR | HADS Anxiety score<br>HADS Depression score | 57<br>57 | 9.6491228<br>5.5964912 | 3.7772402<br>3.8352670 | 3.0000000<br>0 | 9.0000000<br>5.0000000 | 16.0000000<br>12.0000000 | 0<br>0 | 18.0000000<br>19.0000000 |
| managers | 115 | AscoreR<br>DscoreR | HADS Anxiety score<br>HADS Depression score | 115<br>115 | 10.9304348<br>6.8260870 | 4.0969003<br>3.2960925 | 5.0000000<br>1.0000000 | 11.0000000<br>7.0000000 | 18.0000000<br>12.0000000 | 0<br>0 | 21.0000000<br>14.0000000 |
| media and communication | 27 | AscoreR<br>DscoreR | HADS Anxiety score<br>HADS Depression score | 27<br>27 | 10.0370370<br>6.3703704 | 3.7156786<br>3.6810131 | 5.0000000<br>1.0000000 | 10.0000000<br>6.0000000 | 15.0000000<br>14.0000000 | 4.0000000<br>1.0000000 | 17.0000000<br>15.0000000 |
| missing | 33 | AscoreR<br>DscoreR | HADS Anxiety score<br>HADS Depression score | 33<br>33 | 9.4242424<br>6.7575758 | 4.5759583<br>4.5554247 | 0<br>1.0000000 | 9.0000000<br>7.0000000 | 18.0000000<br>16.0000000 | 0<br>0 | 19.0000000<br>18.0000000 |
| other blue collar | 33 | AscoreR<br>DscoreR | HADS Anxiety score<br>HADS Depression score | 33<br>33 | 10.2121212<br>7.3030303 | 4.0755182<br>3.9881453 | 4.0000000<br>0 | 9.0000000<br>7.0000000 | 18.0000000<br>15.0000000 | 3.0000000<br>0 | 19.0000000<br>15.0000000 |
| retail and other sales related occupations | 17 | AscoreR<br>DscoreR | HADS Anxiety score<br>HADS Depression score | 17<br>17 | 11.5882353<br>7.8823529 | 2.5509514<br>4.1515412 | 7.0000000<br>3.0000000 | 12.0000000<br>7.0000000 | 15.0000000<br>18.0000000 | 7.0000000<br>3.0000000 | 15.0000000<br>18.0000000 |
| teachers | 90 | AscoreR<br>DscoreR | HADS Anxiety score<br>HADS Depression score | 90<br>90 | 10.2000000<br>6.5333333 | 4.5250861<br>3.5921262 | 0<br>1.0000000 | 10.5000000<br>6.0000000 | 18.0000000<br>12.0000000 | 0<br>0 | 20.0000000<br>18.0000000 |

#### The MEANS Procedure

**Gender=Male**

| Occupation | N Obs | Variable | Label | N | Mean | Std Dev | 5th Pctl | 50th Pctl | 95th Pctl | Minimum | Maximum |
|---|---|---|---|---|---|---|---|---|---|---|---|
| accountants, financial specialists, and other business operatio | 13 | AscoreR<br>DscoreR | HADS Anxiety score<br>HADS Depression score | 13<br>13 | 6.9230769<br>4.3846154 | 4.1122082<br>2.6937725 | 1.0000000<br>1.0000000 | 6.0000000<br>4.0000000 | 16.0000000<br>9.0000000 | 1.0000000<br>1.0000000 | 16.0000000<br>9.0000000 |
| adminstrative support | 15 | AscoreR<br>DscoreR | HADS Anxiety score<br>HADS Depression score | 15<br>15 | 8.6666667<br>5.7333333 | 3.6968455<br>2.9146592 | 1.0000000<br>1.0000000 | 10.0000000<br>5.0000000 | 13.0000000<br>10.0000000 | 1.0000000<br>1.0000000 | 13.0000000<br>10.0000000 |
| architects and engineers | 15 | AscoreR<br>DscoreR | HADS Anxiety score<br>HADS Depression score | 15<br>15 | 7.7333333<br>4.9333333 | 3.4323392<br>2.7115274 | 1.0000000<br>2.0000000 | 8.0000000<br>4.0000000 | 14.0000000<br>12.0000000 | 1.0000000<br>2.0000000 | 14.0000000<br>12.0000000 |
| artists and performers | 8 | AscoreR<br>DscoreR | HADS Anxiety score<br>HADS Depression score | 8<br>8 | 11.0000000<br>7.6250000 | 5.2915026<br>3.9618719 | 2.0000000<br>0 | 11.5000000<br>7.5000000 | 18.0000000<br>12.0000000 | 2.0000000<br>0 | 18.0000000<br>12.0000000 |
| computer occupations | 14 | AscoreR<br>DscoreR | HADS Anxiety score<br>HADS Depression score | 14<br>14 | 7.3571429<br>4.3571429 | 4.3607893<br>2.2738359 | 1.0000000<br>1.0000000 | 7.0000000<br>4.5000000 | 15.0000000<br>8.0000000 | 1.0000000<br>1.0000000 | 15.0000000<br>8.0000000 |
| counselors, social workers, and community service occupations | 6 | AscoreR<br>DscoreR | HADS Anxiety score<br>HADS Depression score | 6<br>6 | 10.5000000<br>5.0000000 | 6.3166447<br>4.0000000 | 2.0000000<br>1.0000000 | 10.0000000<br>4.0000000 | 18.0000000<br>11.0000000 | 2.0000000<br>1.0000000 | 18.0000000<br>11.0000000 |
| healthcare practitioners and technicians | 19 | AscoreR<br>DscoreR | HADS Anxiety score<br>HADS Depression score | 19<br>19 | 9.4210526<br>6.1052632 | 4.3373800<br>4.3191400 | 4.0000000<br>1.0000000 | 8.0000000<br>6.0000000 | 20.0000000<br>17.0000000 | 4.0000000<br>1.0000000 | 20.0000000<br>17.0000000 |
| laywers and other legal occupations | 9 | AscoreR<br>DscoreR | HADS Anxiety score<br>HADS Depression score | 9<br>9 | 10.5555556<br>6.4444444 | 4.1866189<br>3.3208098 | 5.0000000<br>2.0000000 | 10.0000000<br>6.0000000 | 19.0000000<br>13.0000000 | 5.0000000<br>2.0000000 | 19.0000000<br>13.0000000 |
| librarians and curators | 2 | AscoreR<br>DscoreR | HADS Anxiety score<br>HADS Depression score | 2<br>2 | 10.0000000<br>6.5000000 | 1.4142136<br>0.7071068 | 9.0000000<br>6.0000000 | 10.0000000<br>6.5000000 | 11.0000000<br>7.0000000 | 9.0000000<br>6.0000000 | 11.0000000<br>7.0000000 |

#### The MEANS Procedure

**Gender=Male**

| Occupation | N Obs | Variable | Label | N | Mean | Std Dev | 5th Pctl | 50th Pctl | 95th Pctl | Minimum | Maximum |
|---|---|---|---|---|---|---|---|---|---|---|---|
| life, physical, and social scientists | 21 | AscoreR<br>DscoreR | HADS Anxiety score<br>HADS Depression score | 21<br>21 | 10.0952381<br>5.8095238 | 4.1941002<br>2.9600515 | 4.0000000<br>1.0000000 | 10.0000000<br>5.0000000 | 16.0000000<br>10.0000000 | 1.0000000<br>0 | 18.0000000<br>11.0000000 |
| managers | 47 | AscoreR<br>DscoreR | HADS Anxiety score<br>HADS Depression score | 47<br>47 | 8.1063830<br>5.0425532 | 4.0337384<br>3.4132571 | 2.0000000<br>1.0000000 | 8.0000000<br>5.0000000 | 16.0000000<br>12.0000000 | 0<br>0 | 17.0000000<br>15.0000000 |
| media and communication | 4 | AscoreR<br>DscoreR | HADS Anxiety score<br>HADS Depression score | 4<br>4 | 6.7500000<br>6.7500000 | 4.3493295<br>7.5443135 | 3.0000000<br>2.0000000 | 5.5000000<br>3.5000000 | 13.0000000<br>18.0000000 | 3.0000000<br>2.0000000 | 13.0000000<br>18.0000000 |
| missing | 10 | AscoreR<br>DscoreR | HADS Anxiety score<br>HADS Depression score | 10<br>10 | 9.4000000<br>4.7000000 | 2.9888682<br>2.7507575 | 4.0000000<br>0 | 9.5000000<br>4.0000000 | 14.0000000<br>9.0000000 | 4.0000000<br>0 | 14.0000000<br>9.0000000 |
| other blue collar | 19 | AscoreR<br>DscoreR | HADS Anxiety score<br>HADS Depression score | 19<br>19 | 8.4210526<br>5.5263158 | 5.0806363<br>2.9882812 | 0<br>0 | 8.0000000<br>5.0000000 | 17.0000000<br>12.0000000 | 0<br>0 | 17.0000000<br>12.0000000 |
| retail and other sales related occupations | 7 | AscoreR<br>DscoreR | HADS Anxiety score<br>HADS Depression score | 7<br>7 | 7.0000000<br>7.2857143 | 2.6457513<br>5.2824958 | 4.0000000<br>2.0000000 | 6.0000000<br>7.0000000 | 12.0000000<br>18.0000000 | 4.0000000<br>2.0000000 | 12.0000000<br>18.0000000 |
| teachers | 25 | AscoreR<br>DscoreR | HADS Anxiety score<br>HADS Depression score | 25<br>25 | 6.9600000<br>5.2000000 | 3.3724373<br>4.1432676 | 1.0000000<br>1.0000000 | 8.0000000<br>4.0000000 | 11.0000000<br>13.0000000 | 0<br>0 | 11.0000000<br>16.0000000 |

#### The MEANS Procedure

**Gender=Female**

| Industry | N Obs | Variable | Label | N | Mean | Std Dev | 5th Pctl | 50th Pctl | 95th Pctl | Minimum | Maximum |
|---|---|---|---|---|---|---|---|---|---|---|---|
| accommodation or food services | 14 | AscoreR<br>DscoreR | HADS Anxiety score<br>HADS Depression score | 14<br>14 | 8.2857143<br>6.5000000 | 4.6810349<br>4.5191899 | 0<br>0 | 8.0000000<br>5.5000000 | 16.0000000<br>15.0000000 | 0<br>0 | 16.0000000<br>15.0000000 |
| arts and entertainment | 21 | AscoreR<br>DscoreR | HADS Anxiety score<br>HADS Depression score | 21<br>21 | 10.7142857<br>7.0000000 | 3.6213652<br>2.6457513 | 5.0000000<br>3.0000000 | 11.0000000<br>7.0000000 | 16.0000000<br>11.0000000 | 3.0000000<br>3.0000000 | 16.0000000<br>12.0000000 |
| childcare | 13 | AscoreR<br>DscoreR | HADS Anxiety score<br>HADS Depression score | 13<br>13 | 10.4615385<br>6.8461538 | 5.7679486<br>3.7825510 | 0<br>1.0000000 | 11.0000000<br>7.0000000 | 19.0000000<br>12.0000000 | 0<br>1.0000000 | 19.0000000<br>12.0000000 |
| construction or utilities | 11 | AscoreR<br>DscoreR | HADS Anxiety score<br>HADS Depression score | 11<br>11 | 9.0909091<br>8.2727273 | 3.7270510<br>3.4667249 | 4.0000000<br>3.0000000 | 8.0000000<br>9.0000000 | 15.0000000<br>13.0000000 | 4.0000000<br>3.0000000 | 15.0000000<br>13.0000000 |
| education | 76 | AscoreR<br>DscoreR | HADS Anxiety score<br>HADS Depression score | 76<br>76 | 10.6184211<br>6.7631579 | 4.3816804<br>3.8327307 | 3.0000000<br>1.0000000 | 11.0000000<br>6.0000000 | 19.0000000<br>13.0000000 | 0<br>0 | 20.0000000<br>18.0000000 |
| finance or insurance | 30 | AscoreR<br>DscoreR | HADS Anxiety score<br>HADS Depression score | 30<br>30 | 10.4666667<br>8.3666667 | 4.4001045<br>4.3745607 | 3.0000000<br>1.0000000 | 10.0000000<br>8.0000000 | 18.0000000<br>16.0000000 | 1.0000000<br>1.0000000 | 19.0000000<br>18.0000000 |
| government | 27 | AscoreR<br>DscoreR | HADS Anxiety score<br>HADS Depression score | 27<br>27 | 10.2592593<br>5.9259259 | 4.3287287<br>3.4854194 | 4.0000000<br>2.0000000 | 10.0000000<br>5.0000000 | 16.0000000<br>12.0000000 | 0<br>0 | 16.0000000<br>14.0000000 |
| healthcare | 107 | AscoreR<br>DscoreR | HADS Anxiety score<br>HADS Depression score | 107<br>107 | 10.0186916<br>6.3084112 | 4.0608144<br>3.6352862 | 4.0000000<br>1.0000000 | 10.0000000<br>6.0000000 | 17.0000000<br>12.0000000 | 0<br>0 | 21.0000000<br>20.0000000 |
| higher education | 116 | AscoreR<br>DscoreR | HADS Anxiety score<br>HADS Depression score | 116<br>116 | 9.8620690<br>6.0172414 | 3.7944961<br>3.4690752 | 5.0000000<br>1.0000000 | 10.0000000<br>6.0000000 | 17.0000000<br>12.0000000 | 0<br>0 | 20.0000000<br>16.0000000 |

#### The MEANS Procedure

##### Gender=Female

| Industry | N Obs | Variable | Label | N | Mean | Std Dev | 5th Pctl | 50th Pctl | 95th Pctl | Minimum | Maximum |
|---|---|---|---|---|---|---|---|---|---|---|---|
| legal services | 32 | AscoreR<br>DscoreR | HADS Anxiety score<br>HADS Depression score | 32<br>32 | 9.9375000<br>5.6562500 | 3.9426330<br>2.8916942 | 3.0000000<br>1.0000000 | 10.0000000<br>5.5000000 | 16.0000000<br>10.0000000 | 1.0000000<br>0 | 18.0000000<br>10.0000000 |
| manufacturing | 9 | AscoreR<br>DscoreR | HADS Anxiety score<br>HADS Depression score | 9<br>9 | 9.6666667<br>5.0000000 | 3.0413813<br>3.3911650 | 6.0000000<br>2.0000000 | 9.0000000<br>4.0000000 | 15.0000000<br>10.0000000 | 6.0000000<br>2.0000000 | 15.0000000<br>10.0000000 |
| missing | 26 | AscoreR<br>DscoreR | HADS Anxiety score<br>HADS Depression score | 26<br>26 | 9.3846154<br>7.0000000 | 3.9301595<br>2.4657656 | 5.0000000<br>2.0000000 | 9.0000000<br>7.0000000 | 16.0000000<br>11.0000000 | 0<br>2.0000000 | 21.0000000<br>12.0000000 |
| personal services or repair or other support services | 11 | AscoreR<br>DscoreR | HADS Anxiety score<br>HADS Depression score | 11<br>11 | 12.7272727<br>9.0909091 | 3.1013194<br>3.1130225 | 8.0000000<br>6.0000000 | 14.0000000<br>8.0000000 | 16.0000000<br>15.0000000 | 8.0000000<br>6.0000000 | 16.0000000<br>15.0000000 |
| pharmaceutical | 8 | AscoreR<br>DscoreR | HADS Anxiety score<br>HADS Depression score | 8<br>8 | 9.1250000<br>5.0000000 | 3.7961447<br>3.4226139 | 5.0000000<br>1.0000000 | 8.5000000<br>4.0000000 | 15.0000000<br>12.0000000 | 5.0000000<br>1.0000000 | 15.0000000<br>12.0000000 |
| professional or scientific or engineering or computer services | 36 | AscoreR<br>DscoreR | HADS Anxiety score<br>HADS Depression score | 36<br>36 | 10.0833333<br>5.8888889 | 4.2586047<br>3.7552080 | 4.0000000<br>1.0000000 | 10.0000000<br>6.0000000 | 18.0000000<br>16.0000000 | 2.0000000<br>1.0000000 | 18.0000000<br>17.0000000 |
| publishing or media or other information services | 23 | AscoreR<br>DscoreR | HADS Anxiety score<br>HADS Depression score | 23<br>23 | 10.0869565<br>6.9130435 | 3.5279796<br>3.8602054 | 5.0000000<br>2.0000000 | 9.0000000<br>6.0000000 | 15.0000000<br>14.0000000 | 2.0000000<br>0 | 15.0000000<br>15.0000000 |
| real estate | 6 | AscoreR<br>DscoreR | HADS Anxiety score<br>HADS Depression score | 6<br>6 | 8.5000000<br>4.3333333 | 4.0865633<br>2.5819889 | 5.0000000<br>0 | 7.0000000<br>5.0000000 | 15.0000000<br>7.0000000 | 5.0000000<br>0 | 15.0000000<br>7.0000000 |
| religious or grantmaking or civic or labor organizations | 32 | AscoreR<br>DscoreR | HADS Anxiety score<br>HADS Depression score | 32<br>32 | 9.0937500<br>5.5312500 | 5.1076121<br>3.3599575 | 1.0000000<br>1.0000000 | 8.0000000<br>5.0000000 | 18.0000000<br>13.0000000 | 0<br>0 | 19.0000000<br>14.0000000 |

#### The MEANS Procedure

**Gender=Female**

| Industry | N Obs | Variable | Label | N | Mean | Std Dev | 5th Pctl | 50th Pctl | 95th Pctl | Minimum | Maximum |
|---|---|---|---|---|---|---|---|---|---|---|---|
| retail | 18 | AscoreR<br>DscoreR | HADS Anxiety score<br>HADS Depression score | 18<br>18 | 12.6111111<br>7.8888889 | 2.6377104<br>3.0271106 | 7.0000000<br>3.0000000 | 12.5000000<br>7.5000000 | 19.0000000<br>14.0000000 | 7.0000000<br>3.0000000 | 19.0000000<br>14.0000000 |
| social services | 26 | AscoreR<br>DscoreR | HADS Anxiety score<br>HADS Depression score | 26<br>26 | 9.9615385<br>6.6153846 | 4.5911286<br>4.0306518 | 1.0000000<br>1.0000000 | 9.0000000<br>7.0000000 | 18.0000000<br>14.0000000 | 0<br>0 | 19.0000000<br>14.0000000 |
| telecommunications | 11 | AscoreR<br>DscoreR | HADS Anxiety score<br>HADS Depression score | 11<br>11 | 10.6363636<br>9.0000000 | 4.9854333<br>4.9598387 | 2.0000000<br>1.0000000 | 10.0000000<br>8.0000000 | 19.0000000<br>19.0000000 | 2.0000000<br>1.0000000 | 19.0000000<br>19.0000000 |
| transportation | 8 | AscoreR<br>DscoreR | HADS Anxiety score<br>HADS Depression score | 8<br>8 | 7.6250000<br>4.8750000 | 3.3354160<br>5.8660646 | 4.0000000<br>0 | 7.0000000<br>3.5000000 | 13.0000000<br>18.0000000 | 4.0000000<br>0 | 13.0000000<br>18.0000000 |
| wholesale | 2 | AscoreR<br>DscoreR | HADS Anxiety score<br>HADS Depression score | 2<br>2 | 9.0000000<br>4.0000000 | 1.4142136<br>2.8284271 | 8.0000000<br>2.0000000 | 9.0000000<br>4.0000000 | 10.0000000<br>6.0000000 | 8.0000000<br>2.0000000 | 10.0000000<br>6.0000000 |

**Gender=Male**

| Industry | N Obs | Variable | Label | N | Mean | Std Dev | 5th Pctl | 50th Pctl | 95th Pctl | Minimum | Maximum |
|---|---|---|---|---|---|---|---|---|---|---|---|
| accommodation or food services | 8 | AscoreR<br>DscoreR | HADS Anxiety score<br>HADS Depression score | 8<br>8 | 10.1250000<br>7.0000000 | 3.6815175<br>4.8107024 | 6.0000000<br>3.0000000 | 9.0000000<br>6.0000000 | 17.0000000<br>18.0000000 | 6.0000000<br>3.0000000 | 17.0000000<br>18.0000000 |
| arts and entertainment | 10 | AscoreR<br>DscoreR | HADS Anxiety score<br>HADS Depression score | 10<br>10 | 10.3000000<br>8.8000000 | 3.7133393<br>4.3410188 | 6.0000000<br>3.0000000 | 11.0000000<br>9.5000000 | 18.0000000<br>16.0000000 | 6.0000000<br>3.0000000 | 18.0000000<br>16.0000000 |
| construction or utilities | 15 | AscoreR<br>DscoreR | HADS Anxiety score<br>HADS Depression score | 15<br>15 | 7.5333333<br>4.8666667 | 3.2263794<br>2.4162151 | 1.0000000<br>1.0000000 | 7.0000000<br>4.0000000 | 12.0000000<br>9.0000000 | 1.0000000<br>1.0000000 | 12.0000000<br>9.0000000 |
| education | 9 | AscoreR<br>DscoreR | HADS Anxiety score<br>HADS Depression score | 9<br>9 | 8.5555556<br>4.8888889 | 2.8771128<br>3.1001792 | 4.0000000<br>0 | 9.0000000<br>4.0000000 | 13.0000000<br>10.0000000 | 4.0000000<br>0 | 13.0000000<br>10.0000000 |

#### The MEANS Procedure

**Gender=Male**

| Industry | N Obs | Variable | Label | N | Mean | Std Dev | 5th Pctl | 50th Pctl | 95th Pctl | Minimum | Maximum |
|---|---|---|---|---|---|---|---|---|---|---|---|
| finance or insurance | 12 | AscoreR<br>DscoreR | HADS Anxiety score<br>HADS Depression score | 12<br>12 | 6.8333333<br>4.0000000 | 3.7132033<br>2.7961012 | 1.0000000<br>1.0000000 | 6.0000000<br>3.0000000 | 13.0000000<br>9.0000000 | 1.0000000<br>1.0000000 | 13.0000000<br>9.0000000 |
| government | 12 | AscoreR<br>DscoreR | HADS Anxiety score<br>HADS Depression score | 12<br>12 | 5.2500000<br>3.7500000 | 4.0704032<br>3.4673805 | 0<br>0 | 5.0000000<br>2.5000000 | 13.0000000<br>10.0000000 | 0<br>0 | 13.0000000<br>10.0000000 |
| healthcare | 30 | AscoreR<br>DscoreR | HADS Anxiety score<br>HADS Depression score | 30<br>30 | 10.1333333<br>5.8333333 | 4.5541443<br>3.9047569 | 4.0000000<br>1.0000000 | 9.0000000<br>6.0000000 | 18.0000000<br>14.0000000 | 3.0000000<br>1.0000000 | 20.0000000<br>17.0000000 |
| higher education | 36 | AscoreR<br>DscoreR | HADS Anxiety score<br>HADS Depression score | 36<br>36 | 7.9722222<br>5.1666667 | 3.8580981<br>3.1396087 | 1.0000000<br>1.0000000 | 8.0000000<br>5.0000000 | 14.0000000<br>12.0000000 | 0<br>1.0000000 | 18.0000000<br>13.0000000 |
| legal services | 8 | AscoreR<br>DscoreR | HADS Anxiety score<br>HADS Depression score | 8<br>8 | 10.2500000<br>6.1250000 | 4.3670847<br>3.3990545 | 5.0000000<br>2.0000000 | 10.0000000<br>5.5000000 | 19.0000000<br>13.0000000 | 5.0000000<br>2.0000000 | 19.0000000<br>13.0000000 |
| manufacturing | 5 | AscoreR<br>DscoreR | HADS Anxiety score<br>HADS Depression score | 5<br>5 | 5.4000000<br>4.2000000 | 4.4497191<br>4.4944410 | 0<br>1.0000000 | 4.0000000<br>2.0000000 | 12.0000000<br>12.0000000 | 0<br>1.0000000 | 12.0000000<br>12.0000000 |
| missing | 10 | AscoreR<br>DscoreR | HADS Anxiety score<br>HADS Depression score | 10<br>10 | 6.8000000<br>4.9000000 | 3.4576807<br>1.6633300 | 2.0000000<br>3.0000000 | 7.0000000<br>4.5000000 | 14.0000000<br>8.0000000 | 2.0000000<br>3.0000000 | 14.0000000<br>8.0000000 |
| personal services or repair or other support services | 3 | AscoreR<br>DscoreR | HADS Anxiety score<br>HADS Depression score | 3<br>3 | 9.3333333<br>3.6666667 | 6.8068593<br>3.2145503 | 4.0000000<br>0 | 7.0000000<br>5.0000000 | 17.0000000<br>6.0000000 | 4.0000000<br>0 | 17.0000000<br>6.0000000 |
| pharmaceutical | 8 | AscoreR<br>DscoreR | HADS Anxiety score<br>HADS Depression score | 8<br>8 | 8.7500000<br>5.0000000 | 4.8916839<br>3.0237158 | 1.0000000<br>0 | 9.5000000<br>5.0000000 | 16.0000000<br>9.0000000 | 1.0000000<br>0 | 16.0000000<br>9.0000000 |

#### The MEANS Procedure

**Gender=Male**

| Industry | N Obs | Variable | Label | N | Mean | Std Dev | 5th Pctl | 50th Pctl | 95th Pctl | Minimum | Maximum |
|---|---|---|---|---|---|---|---|---|---|---|---|
| professional or scientific or engineering or computer services | 26 | AscoreR<br>DscoreR | HADS Anxiety score<br>HADS Depression score | 26<br>26 | 8.1538462<br>5.0769231 | 3.4950514<br>2.5443754 | 4.0000000<br>1.0000000 | 8.0000000<br>5.0000000 | 14.0000000<br>9.0000000 | 2.0000000<br>0 | 15.0000000<br>10.0000000 |
| publishing or media or other information services | 5 | AscoreR<br>DscoreR | HADS Anxiety score<br>HADS Depression score | 5<br>5 | 11.4000000<br>9.6000000 | 2.7018512<br>6.1886994 | 8.0000000<br>3.0000000 | 13.0000000<br>7.0000000 | 14.0000000<br>18.0000000 | 8.0000000<br>3.0000000 | 14.0000000<br>18.0000000 |
| real estate | 3 | AscoreR<br>DscoreR | HADS Anxiety score<br>HADS Depression score | 3<br>3 | 11.0000000<br>9.3333333 | 2.6457513<br>5.5075705 | 8.0000000<br>4.0000000 | 12.0000000<br>9.0000000 | 13.0000000<br>15.0000000 | 8.0000000<br>4.0000000 | 13.0000000<br>15.0000000 |
| religious or grantmaking or civic or labor organizations | 7 | AscoreR<br>DscoreR | HADS Anxiety score<br>HADS Depression score | 7<br>7 | 7.5714286<br>5.2857143 | 3.1014590<br>3.1997024 | 2.0000000<br>2.0000000 | 9.0000000<br>4.0000000 | 11.0000000<br>12.0000000 | 2.0000000<br>2.0000000 | 11.0000000<br>12.0000000 |
| retail | 11 | AscoreR<br>DscoreR | HADS Anxiety score<br>HADS Depression score | 11<br>11 | 9.0000000<br>4.9090909 | 5.7445626<br>3.3303017 | 1.0000000<br>1.0000000 | 8.0000000<br>5.0000000 | 17.0000000<br>12.0000000 | 1.0000000<br>1.0000000 | 17.0000000<br>12.0000000 |
| social services | 8 | AscoreR<br>DscoreR | HADS Anxiety score<br>HADS Depression score | 8<br>8 | 11.0000000<br>6.5000000 | 4.6291005<br>3.7416574 | 4.0000000<br>0 | 11.0000000<br>6.5000000 | 17.0000000<br>11.0000000 | 4.0000000<br>0 | 17.0000000<br>11.0000000 |
| telecommunications | 2 | AscoreR<br>DscoreR | HADS Anxiety score<br>HADS Depression score | 2<br>2 | 5.5000000<br>3.5000000 | 2.1213203<br>0.7071068 | 4.0000000<br>3.0000000 | 5.5000000<br>3.5000000 | 7.0000000<br>4.0000000 | 4.0000000<br>3.0000000 | 7.0000000<br>4.0000000 |
| transportation | 4 | AscoreR<br>DscoreR | HADS Anxiety score<br>HADS Depression score | 4<br>4 | 6.7500000<br>4.7500000 | 3.7749172<br>2.5000000 | 4.0000000<br>2.0000000 | 5.5000000<br>4.5000000 | 12.0000000<br>8.0000000 | 4.0000000<br>2.0000000 | 12.0000000<br>8.0000000 |
| wholesale | 2 | AscoreR<br>DscoreR | HADS Anxiety score<br>HADS Depression score | 2<br>2 | 8.0000000<br>6.5000000 | 8.4852814<br>0.7071068 | 2.0000000<br>6.0000000 | 8.0000000<br>6.5000000 | 14.0000000<br>7.0000000 | 2.0000000<br>6.0000000 | 14.0000000<br>7.0000000 |

#### The MEANS Procedure

##### Gender=Female

| Work arrangement changed by Stay at home order March 23 | N Obs | Variable | Label | N | Mean | Std Dev | 5th Pctl | 50th Pctl | 95th Pctl | Minimum | Maximum |
|---|---|---|---|---|---|---|---|---|---|---|---|
| no | 173 | AscoreR<br>DscoreR | HADS Anxiety score<br>HADS Depression score | 173<br>173 | 9.8612717<br>6.6242775 | 4.3779777<br>3.9580489 | 2.0000000<br>1.0000000 | 10.0000000<br>7.0000000 | 18.0000000<br>14.0000000 | 0<br>0 | 21.0000000<br>20.0000000 |
| yes | 489 | AscoreR<br>DscoreR | HADS Anxiety score<br>HADS Depression score | 489<br>489 | 10.1308793<br>6.4335378 | 4.0477741<br>3.5770472 | 4.0000000<br>1.0000000 | 10.0000000<br>6.0000000 | 17.0000000<br>13.0000000 | 0<br>0 | 20.0000000<br>19.0000000 |
| missing | 1 | AscoreR<br>DscoreR | HADS Anxiety score<br>HADS Depression score | 1<br>1 | 12.0000000<br>6.0000000 | .<br>. | 12.0000000<br>6.0000000 | 12.0000000<br>6.0000000 | 12.0000000<br>6.0000000 | 12.0000000<br>6.0000000 | 12.0000000<br>6.0000000 |

##### Gender=Male

| Work arrangement changed by Stay at home order March 23 | N Obs | Variable | Label | N | Mean | Std Dev | 5th Pctl | 50th Pctl | 95th Pctl | Minimum | Maximum |
|---|---|---|---|---|---|---|---|---|---|---|---|
| no | 79 | AscoreR<br>DscoreR | HADS Anxiety score<br>HADS Depression score | 79<br>79 | 7.8227848<br>5.1139241 | 4.3345940<br>3.2105090 | 1.0000000<br>1.0000000 | 7.0000000<br>5.0000000 | 17.0000000<br>12.0000000 | 0<br>0 | 20.0000000<br>13.0000000 |
| yes | 155 | AscoreR<br>DscoreR | HADS Anxiety score<br>HADS Depression score | 155<br>155 | 8.7935484<br>5.6193548 | 4.0559348<br>3.6096389 | 2.0000000<br>1.0000000 | 9.0000000<br>5.0000000 | 16.0000000<br>13.0000000 | 0<br>0 | 18.0000000<br>18.0000000 |

#### The MEANS Procedure

##### Gender=Female

| Were unwell for two or more consecutive days (whether or not worked) | N Obs | Variable | Label | N | Mean | Std Dev | 5th Pctl | 50th Pctl | 95th Pctl | Minimum | Maximum |
|---|---|---|---|---|---|---|---|---|---|---|---|
| no | 498 | AscoreR<br>DscoreR | HADS Anxiety score<br>HADS Depression score | 498<br>498 | 9.8755020<br>6.3132530 | 4.2403355<br>3.7419619 | 3.0000000<br>1.0000000 | 10.0000000<br>6.0000000 | 18.0000000<br>13.0000000 | 0<br>0 | 21.0000000<br>20.0000000 |
| yes | 164 | AscoreR<br>DscoreR | HADS Anxiety score<br>HADS Depression score | 164<br>164 | 10.6585366<br>7.0000000 | 3.7374560<br>3.4374338 | 5.0000000<br>2.0000000 | 10.0000000<br>7.0000000 | 16.0000000<br>14.0000000 | 0<br>0 | 20.0000000<br>19.0000000 |
| missing | 1 | AscoreR<br>DscoreR | HADS Anxiety score<br>HADS Depression score | 1<br>1 | 6.0000000<br>6.0000000 | .<br>. | 6.0000000<br>6.0000000 | 6.0000000<br>6.0000000 | 6.0000000<br>6.0000000 | 6.0000000<br>6.0000000 | 6.0000000<br>6.0000000 |

##### Gender=Male

| Were unwell for two or more consecutive days (whether or not worked) | N Obs | Variable | Label | N | Mean | Std Dev | 5th Pctl | 50th Pctl | 95th Pctl | Minimum | Maximum |
|---|---|---|---|---|---|---|---|---|---|---|---|
| no | 188 | AscoreR<br>DscoreR | HADS Anxiety score<br>HADS Depression score | 188<br>188 | 8.1968085<br>5.2446809 | 4.3174302<br>3.4754638 | 1.0000000<br>1.0000000 | 8.0000000<br>5.0000000 | 16.0000000<br>12.0000000 | 0<br>0 | 20.0000000<br>18.0000000 |
| yes | 45 | AscoreR<br>DscoreR | HADS Anxiety score<br>HADS Depression score | 45<br>45 | 9.6666667<br>6.3777778 | 3.2752516<br>3.3931005 | 6.0000000<br>2.0000000 | 9.0000000<br>6.0000000 | 17.0000000<br>12.0000000 | 5.0000000<br>1.0000000 | 18.0000000<br>18.0000000 |
| missing | 1 | AscoreR<br>DscoreR | HADS Anxiety score<br>HADS Depression score | 1<br>1 | 5.0000000<br>2.0000000 | .<br>. | 5.0000000<br>2.0000000 | 5.0000000<br>2.0000000 | 5.0000000<br>2.0000000 | 5.0000000<br>2.0000000 | 5.0000000<br>2.0000000 |

### All who had a job

#### The MEANS Procedure

##### Gender=Female

| Believe may have been infected | N Obs | Variable | Label | N | Mean | Std Dev | 5th Pctl | 50th Pctl | 95th Pctl | Minimum | Maximum |
|---|---|---|---|---|---|---|---|---|---|---|---|
| no | 475 | AscoreR | HADS Anxiety score | 475 | 9.8821053 | 4.1316413 | 3.0000000 | 10.0000000 | 16.0000000 | 0 | 21.0000000 |
| | | DscoreR | HADS Depression score | 475 | 6.2926316 | 3.6711684 | 1.0000000 | 6.0000000 | 13.0000000 | 0 | 20.0000000 |
| yes | 59 | AscoreR | HADS Anxiety score | 59 | 11.1525424 | 4.5893939 | 4.0000000 | 11.0000000 | 19.0000000 | 0 | 20.0000000 |
| | | DscoreR | HADS Depression score | 59 | 7.3898305 | 3.8011657 | 1.0000000 | 7.0000000 | 14.0000000 | 0 | 16.0000000 |
| missing | 129 | AscoreR | HADS Anxiety score | 129 | 10.2325581 | 3.8558228 | 5.0000000 | 10.0000000 | 16.0000000 | 0 | 20.0000000 |
| | | DscoreR | HADS Depression score | 129 | 6.7674419 | 3.5805718 | 1.0000000 | 7.0000000 | 14.0000000 | 0 | 18.0000000 |

##### Gender=Male

| Believe may have been infected | N Obs | Variable | Label | N | Mean | Std Dev | 5th Pctl | 50th Pctl | 95th Pctl | Minimum | Maximum |
|---|---|---|---|---|---|---|---|---|---|---|---|
| no | 175 | AscoreR | HADS Anxiety score | 175 | 8.0685714 | 4.1420032 | 2.0000000 | 8.0000000 | 16.0000000 | 0 | 20.0000000 |
| | | DscoreR | HADS Depression score | 175 | 5.3257143 | 3.5738790 | 1.0000000 | 5.0000000 | 12.0000000 | 0 | 18.0000000 |
| yes | 20 | AscoreR | HADS Anxiety score | 20 | 9.1500000 | 4.8587956 | 2.0000000 | 8.0000000 | 17.0000000 | 1.0000000 | 17.0000000 |
| | | DscoreR | HADS Depression score | 20 | 5.2000000 | 2.6675437 | 1.0000000 | 4.5000000 | 9.0000000 | 0 | 9.0000000 |
| missing | 39 | AscoreR | HADS Anxiety score | 39 | 9.8974359 | 3.6186278 | 4.0000000 | 10.0000000 | 17.0000000 | 1.0000000 | 18.0000000 |
| | | DscoreR | HADS Depression score | 39 | 6.1282051 | 3.4195990 | 1.0000000 | 5.0000000 | 12.0000000 | 1.0000000 | 14.0000000 |

#### The MEANS Procedure

**Gender=Female**

| Self-rated health compared to others | N Obs | Variable | Label | N | Mean | Std Dev | 5th Pctl | 50th Pctl | 95th Pctl | Minimum | Maximum |
|---|---|---|---|---|---|---|---|---|---|---|---|
| good to excellent | 594 | AscoreR<br>DscoreR | HADS Anxiety score<br>HADS Depression score | 594<br>594 | 9.8552189<br>6.2777778 | 4.1129805<br>3.6256456 | 3.0000000<br>1.0000000 | 10.0000000<br>6.0000000 | 17.0000000<br>12.0000000 | 0<br>0 | 21.0000000<br>20.0000000 |
| poor or fair | 69 | AscoreR<br>DscoreR | HADS Anxiety score<br>HADS Depression score | 69<br>69 | 11.8550725<br>8.2463768 | 3.8854001<br>3.6558004 | 7.0000000<br>2.0000000 | 12.0000000<br>8.0000000 | 19.0000000<br>14.0000000 | 5.0000000<br>1.0000000 | 20.0000000<br>16.0000000 |

**Gender=Male**

| Self-rated health compared to others | N Obs | Variable | Label | N | Mean | Std Dev | 5th Pctl | 50th Pctl | 95th Pctl | Minimum | Maximum |
|---|---|---|---|---|---|---|---|---|---|---|---|
| good to excellent | 211 | AscoreR<br>DscoreR | HADS Anxiety score<br>HADS Depression score | 211<br>211 | 8.2843602<br>5.1658768 | 4.1969027<br>3.1631768 | 2.0000000<br>1.0000000 | 8.0000000<br>5.0000000 | 16.0000000<br>11.0000000 | 0<br>0 | 20.0000000<br>18.0000000 |
| poor or fair | 23 | AscoreR<br>DscoreR | HADS Anxiety score<br>HADS Depression score | 23<br>23 | 10.1304348<br>8.0434783 | 3.5586559<br>5.0043459 | 4.0000000<br>1.0000000 | 10.0000000<br>6.0000000 | 16.0000000<br>17.0000000 | 4.0000000<br>0 | 17.0000000<br>18.0000000 |

#### The MEANS Procedure

##### Gender=Female

| Self-rated health | N Obs | Variable | Label | N | Mean | Std Dev | 5th Pctl | 50th Pctl | 95th Pctl | Minimum | Maximum |
|---|---|---|---|---|---|---|---|---|---|---|---|
| good to excellent | 633 | AscoreR<br>DscoreR | HADS Anxiety score<br>HADS Depression score | 633<br>633 | 9.9921011<br>6.4312796 | 4.1121464<br>3.6507457 | 4.0000000<br>1.0000000 | 10.0000000<br>6.0000000 | 17.0000000<br>13.0000000 | 0<br>0 | 21.0000000<br>20.0000000 |
| poor or fair | 30 | AscoreR<br>DscoreR | HADS Anxiety score<br>HADS Depression score | 30<br>30 | 11.5666667<br>7.5666667 | 4.3445037<br>4.0826237 | 7.0000000<br>2.0000000 | 11.0000000<br>6.0000000 | 20.0000000<br>15.0000000 | 7.0000000<br>1.0000000 | 20.0000000<br>16.0000000 |

##### Gender=Male

| Self-rated health | N Obs | Variable | Label | N | Mean | Std Dev | 5th Pctl | 50th Pctl | 95th Pctl | Minimum | Maximum |
|---|---|---|---|---|---|---|---|---|---|---|---|
| good to excellent | 228 | AscoreR<br>DscoreR | HADS Anxiety score<br>HADS Depression score | 228<br>228 | 8.3991228<br>5.3026316 | 4.1378663<br>3.3013937 | 2.0000000<br>1.0000000 | 8.0000000<br>5.0000000 | 16.0000000<br>12.0000000 | 0<br>0 | 20.0000000<br>18.0000000 |
| poor or fair | 6 | AscoreR<br>DscoreR | HADS Anxiety score<br>HADS Depression score | 6<br>6 | 11.0000000<br>11.0000000 | 4.9396356<br>5.6568542 | 5.0000000<br>2.0000000 | 11.0000000<br>12.0000000 | 17.0000000<br>17.0000000 | 5.0000000<br>2.0000000 | 17.0000000<br>17.0000000 |

#### The MEANS Procedure

##### Gender=Female

| Phase of restrictions | N Obs | Variable | Label | N | Mean | Std Dev | 5th Pctl | 50th Pctl | 95th Pctl | Minimum | Maximum |
|---|---|---|---|---|---|---|---|---|---|---|---|
| red | 370 | AscoreR<br>DscoreR | HADS Anxiety score<br>HADS Depression score | 370<br>370 | 9.8621622<br>6.3081081 | 4.2195731<br>3.6234989 | 3.0000000<br>1.0000000 | 10.0000000<br>6.0000000 | 16.0000000<br>13.0000000 | 0<br>0 | 21.0000000<br>20.0000000 |
| yellow | 293 | AscoreR<br>DscoreR | HADS Anxiety score<br>HADS Depression score | 293<br>293 | 10.3174061<br>6.7030717 | 4.0126005<br>3.7348632 | 4.0000000<br>1.0000000 | 10.0000000<br>7.0000000 | 18.0000000<br>13.0000000 | 0<br>0 | 20.0000000<br>18.0000000 |

##### Gender=Male

| Phase of restrictions | N Obs | Variable | Label | N | Mean | Std Dev | 5th Pctl | 50th Pctl | 95th Pctl | Minimum | Maximum |
|---|---|---|---|---|---|---|---|---|---|---|---|
| red | 146 | AscoreR<br>DscoreR | HADS Anxiety score<br>HADS Depression score | 146<br>146 | 8.2739726<br>5.2054795 | 3.9401042<br>3.3629171 | 2.0000000<br>1.0000000 | 8.0000000<br>5.0000000 | 14.0000000<br>12.0000000 | 0<br>0 | 19.0000000<br>18.0000000 |
| yellow | 88 | AscoreR<br>DscoreR | HADS Anxiety score<br>HADS Depression score | 88<br>88 | 8.7840909<br>5.8522727 | 4.5269280<br>3.6531824 | 2.0000000<br>1.0000000 | 8.0000000<br>5.0000000 | 17.0000000<br>12.0000000 | 0<br>0 | 20.0000000<br>18.0000000 |

### Only those who reported change in work since first case

#### The MEANS Procedure

##### Gender=Female

| Lost job | N Obs | Variable | Label | N | Mean | Std Dev | 5th Pctl | 50th Pctl | 95th Pctl | Minimum | Maximum |
|---|---|---|---|---|---|---|---|---|---|---|---|
| no | 436 | AscoreR | HADS Anxiety score | 436 | 10.1055046 | 4.0032018 | 4.0000000 | 10.0000000 | 17.0000000 | 0 | 20.0000000 |
| | | DscoreR | HADS Depression score | 436 | 6.3967890 | 3.5668833 | 1.0000000 | 6.0000000 | 12.0000000 | 0 | 19.0000000 |
| yes | 53 | AscoreR | HADS Anxiety score | 53 | 10.3396226 | 4.4330203 | 3.0000000 | 10.0000000 | 18.0000000 | 3.0000000 | 20.0000000 |
| | | DscoreR | HADS Depression score | 53 | 6.7358491 | 3.6802537 | 0 | 7.0000000 | 14.0000000 | 0 | 15.0000000 |

##### Gender=Male

| Lost job | N Obs | Variable | Label | N | Mean | Std Dev | 5th Pctl | 50th Pctl | 95th Pctl | Minimum | Maximum |
|---|---|---|---|---|---|---|---|---|---|---|---|
| no | 141 | AscoreR | HADS Anxiety score | 141 | 8.5602837 | 3.9358586 | 2.0000000 | 9.0000000 | 15.0000000 | 0 | 18.0000000 |
| | | DscoreR | HADS Depression score | 141 | 5.2978723 | 3.4116286 | 1.0000000 | 5.0000000 | 12.0000000 | 0 | 18.0000000 |
| yes | 14 | AscoreR | HADS Anxiety score | 14 | 11.1428571 | 4.6385864 | 4.0000000 | 11.5000000 | 18.0000000 | 4.0000000 | 18.0000000 |
| | | DscoreR | HADS Depression score | 14 | 8.8571429 | 4.0735005 | 4.0000000 | 8.0000000 | 18.0000000 | 4.0000000 | 18.0000000 |

### Only those who reported change in work since first case

#### The MEANS Procedure

##### Gender=Female

| Applied for unemployement | N Obs | Variable | Label | N | Mean | Std Dev | 5th Pctl | 50th Pctl | 95th Pctl | Minimum | Maximum |
|---|---|---|---|---|---|---|---|---|---|---|---|
| no | 14 | AscoreR<br>DscoreR | HADS Anxiety score<br>HADS Depression score | 14<br>14 | 10.8571429<br>7.9285714 | 5.4187302<br>3.4073289 | 3.0000000<br>3.0000000 | 10.0000000<br>7.0000000 | 20.0000000<br>15.0000000 | 3.0000000<br>3.0000000 | 20.0000000<br>15.0000000 |
| yes | 38 | AscoreR<br>DscoreR | HADS Anxiety score<br>HADS Depression score | 38<br>38 | 10.2894737<br>6.2105263 | 4.0530795<br>3.7210714 | 4.0000000<br>0 | 10.0000000<br>6.0000000 | 18.0000000<br>14.0000000 | 3.0000000<br>0 | 19.0000000<br>15.0000000 |
| missing | 437 | AscoreR<br>DscoreR | HADS Anxiety score<br>HADS Depression score | 437<br>437 | 10.0938215<br>6.4050343 | 4.0060600<br>3.5669575 | 4.0000000<br>1.0000000 | 10.0000000<br>6.0000000 | 17.0000000<br>12.0000000 | 0<br>0 | 20.0000000<br>19.0000000 |

##### Gender=Male

| Applied for unemployement | N Obs | Variable | Label | N | Mean | Std Dev | 5th Pctl | 50th Pctl | 95th Pctl | Minimum | Maximum |
|---|---|---|---|---|---|---|---|---|---|---|---|
| no | 4 | AscoreR<br>DscoreR | HADS Anxiety score<br>HADS Depression score | 4<br>4 | 13.0000000<br>11.7500000 | 2.8284271<br>3.0956959 | 11.0000000<br>9.0000000 | 12.0000000<br>11.0000000 | 17.0000000<br>16.0000000 | 11.0000000<br>9.0000000 | 17.0000000<br>16.0000000 |
| yes | 10 | AscoreR<br>DscoreR | HADS Anxiety score<br>HADS Depression score | 10<br>10 | 10.4000000<br>7.7000000 | 5.1251016<br>3.9454615 | 4.0000000<br>4.0000000 | 11.5000000<br>6.5000000 | 18.0000000<br>18.0000000 | 4.0000000<br>4.0000000 | 18.0000000<br>18.0000000 |
| missing | 141 | AscoreR<br>DscoreR | HADS Anxiety score<br>HADS Depression score | 141<br>141 | 8.5602837<br>5.2978723 | 3.9358586<br>3.4116286 | 2.0000000<br>1.0000000 | 9.0000000<br>5.0000000 | 15.0000000<br>12.0000000 | 0<br>0 | 18.0000000<br>18.0000000 |

### Only those who reported change in work since first case

#### The MEANS Procedure

##### Gender=Female

| hrschange | N Obs | Variable | Label | N | Mean | Std Dev | 5th Pctl | 50th Pctl | 95th Pctl | Minimum | Maximum |
|---|---|---|---|---|---|---|---|---|---|---|---|
| about the same | 214 | AscoreR<br>DscoreR | HADS Anxiety score<br>HADS Depression score | 214<br>214 | 9.9906542<br>6.0841121 | 3.7840667<br>3.5830102 | 4.0000000<br>1.0000000 | 10.0000000<br>6.0000000 | 18.0000000<br>12.0000000 | 0<br>0 | 18.0000000<br>19.0000000 |
| more | 83 | AscoreR<br>DscoreR | HADS Anxiety score<br>HADS Depression score | 83<br>83 | 11.1807229<br>6.8192771 | 4.1530024<br>3.2578033 | 5.0000000<br>2.0000000 | 12.0000000<br>7.0000000 | 16.0000000<br>12.0000000 | 0<br>1.0000000 | 20.0000000<br>18.0000000 |
| less | 138 | AscoreR<br>DscoreR | HADS Anxiety score<br>HADS Depression score | 138<br>138 | 9.6231884<br>6.6449275 | 4.1613496<br>3.7040139 | 3.0000000<br>1.0000000 | 9.5000000<br>6.0000000 | 17.0000000<br>13.0000000 | 0<br>0 | 20.0000000<br>18.0000000 |
| missing | 1 | AscoreR<br>DscoreR | HADS Anxiety score<br>HADS Depression score | 1<br>1 | 12.0000000<br>4.0000000 | .<br>. | 12.0000000<br>4.0000000 | 12.0000000<br>4.0000000 | 12.0000000<br>4.0000000 | 12.0000000<br>4.0000000 | 12.0000000<br>4.0000000 |
| lost work | 53 | AscoreR<br>DscoreR | HADS Anxiety score<br>HADS Depression score | 53<br>53 | 10.3396226<br>6.7358491 | 4.4330203<br>3.6802537 | 3.0000000<br>0 | 10.0000000<br>7.0000000 | 18.0000000<br>14.0000000 | 3.0000000<br>0 | 20.0000000<br>15.0000000 |

##### Gender=Male

| hrschange | N Obs | Variable | Label | N | Mean | Std Dev | 5th Pctl | 50th Pctl | 95th Pctl | Minimum | Maximum |
|---|---|---|---|---|---|---|---|---|---|---|---|
| about the same | 69 | AscoreR<br>DscoreR | HADS Anxiety score<br>HADS Depression score | 69<br>69 | 8.3043478<br>4.7971014 | 3.8551125<br>3.1369698 | 1.0000000<br>1.0000000 | 8.0000000<br>4.0000000 | 14.0000000<br>10.0000000 | 0<br>0 | 16.0000000<br>17.0000000 |
| more | 30 | AscoreR<br>DscoreR | HADS Anxiety score<br>HADS Depression score | 30<br>30 | 9.3666667<br>5.3333333 | 4.2384393<br>2.6695387 | 2.0000000<br>1.0000000 | 9.0000000<br>5.0000000 | 17.0000000<br>9.0000000 | 2.0000000<br>1.0000000 | 18.0000000<br>14.0000000 |
| less | 42 | AscoreR<br>DscoreR | HADS Anxiety score<br>HADS Depression score | 42<br>42 | 8.4047619<br>6.0952381 | 3.8639012<br>4.1602622 | 2.0000000<br>1.0000000 | 8.0000000<br>5.5000000 | 15.0000000<br>13.0000000 | 1.0000000<br>0 | 16.0000000<br>18.0000000 |
| lost work | 14 | AscoreR<br>DscoreR | HADS Anxiety score<br>HADS Depression score | 14<br>14 | 11.1428571<br>8.8571429 | 4.6385864<br>4.0735005 | 4.0000000<br>4.0000000 | 11.5000000<br>8.0000000 | 18.0000000<br>18.0000000 | 4.0000000<br>4.0000000 | 18.0000000<br>18.0000000 |

### Only those who reported change in work since first case

#### The MEANS Procedure

##### Gender=Female

| Start or substantially increase telecommuting | N Obs | Variable | Label | N | Mean | Std Dev | 5th Pctl | 50th Pctl | 95th Pctl | Minimum | Maximum |
|---|---|---|---|---|---|---|---|---|---|---|---|
| no | 55 | AscoreR<br>DscoreR | HADS Anxiety score<br>HADS Depression score | 55<br>55 | 10.4727273<br>6.6545455 | 4.4715336<br>3.7918928 | 3.0000000<br>1.0000000 | 10.0000000<br>6.0000000 | 19.0000000<br>14.0000000 | 1.0000000<br>0 | 19.0000000<br>18.0000000 |
| yes | 342 | AscoreR<br>DscoreR | HADS Anxiety score<br>HADS Depression score | 342<br>342 | 10.0321637<br>6.4269006 | 3.9007655<br>3.5035267 | 4.0000000<br>1.0000000 | 10.0000000<br>6.0000000 | 16.0000000<br>12.0000000 | 0<br>0 | 20.0000000<br>19.0000000 |
| missing | 92 | AscoreR<br>DscoreR | HADS Anxiety score<br>HADS Depression score | 92<br>92 | 10.2934783<br>6.3260870 | 4.3413120<br>3.7478454 | 3.0000000<br>0 | 10.0000000<br>6.0000000 | 18.0000000<br>14.0000000 | 0<br>0 | 20.0000000<br>15.0000000 |

##### Gender=Male

| Start or substantially increase telecommuting | N Obs | Variable | Label | N | Mean | Std Dev | 5th Pctl | 50th Pctl | 95th Pctl | Minimum | Maximum |
|---|---|---|---|---|---|---|---|---|---|---|---|
| no | 18 | AscoreR<br>DscoreR | HADS Anxiety score<br>HADS Depression score | 18<br>18 | 9.1111111<br>5.9444444 | 3.4622143<br>3.7333508 | 2.0000000<br>2.0000000 | 9.5000000<br>5.0000000 | 16.0000000<br>18.0000000 | 2.0000000<br>2.0000000 | 16.0000000<br>18.0000000 |
| yes | 105 | AscoreR<br>DscoreR | HADS Anxiety score<br>HADS Depression score | 105<br>105 | 8.7714286<br>4.9904762 | 3.8586993<br>3.1240237 | 2.0000000<br>1.0000000 | 9.0000000<br>4.0000000 | 15.0000000<br>12.0000000 | 1.0000000<br>0 | 18.0000000<br>14.0000000 |
| missing | 32 | AscoreR<br>DscoreR | HADS Anxiety score<br>HADS Depression score | 32<br>32 | 8.6875000<br>7.5000000 | 5.0124844<br>4.3920750 | 0<br>0 | 9.0000000<br>6.0000000 | 17.0000000<br>17.0000000 | 0<br>0 | 18.0000000<br>18.0000000 |

### Only those who reported change in work since first case

#### The MEANS Procedure

##### Gender=Female

| Concerned about returm to work | N Obs | Variable | Label | N | Mean | Std Dev | 5th Pctl | 50th Pctl | 95th Pctl | Minimum | Maximum |
|---|---|---|---|---|---|---|---|---|---|---|---|
| no | 153 | AscoreR<br>DscoreR | HADS Anxiety score<br>HADS Depression score | 153<br>153 | 9.1307190<br>5.8300654 | 4.0679958<br>3.4465674 | 1.0000000<br>1.0000000 | 9.0000000<br>5.0000000 | 16.0000000<br>12.0000000 | 0<br>0 | 20.0000000<br>19.0000000 |
| yes | 336 | AscoreR<br>DscoreR | HADS Anxiety score<br>HADS Depression score | 336<br>336 | 10.5863095<br>6.7083333 | 3.9615680<br>3.6065515 | 5.0000000<br>1.0000000 | 11.0000000<br>7.0000000 | 18.0000000<br>13.0000000 | 0<br>0 | 20.0000000<br>18.0000000 |

##### Gender=Male

| Concerned about returm to work | N Obs | Variable | Label | N | Mean | Std Dev | 5th Pctl | 50th Pctl | 95th Pctl | Minimum | Maximum |
|---|---|---|---|---|---|---|---|---|---|---|---|
| no | 67 | AscoreR<br>DscoreR | HADS Anxiety score<br>HADS Depression score | 67<br>67 | 7.5820896<br>4.8059701 | 4.2503093<br>3.5085990 | 1.0000000<br>1.0000000 | 7.0000000<br>4.0000000 | 16.0000000<br>10.0000000 | 0<br>0 | 17.0000000<br>18.0000000 |
| yes | 88 | AscoreR<br>DscoreR | HADS Anxiety score<br>HADS Depression score | 88<br>88 | 9.7159091<br>6.2386364 | 3.6638930<br>3.5815437 | 4.0000000<br>2.0000000 | 10.0000000<br>6.0000000 | 16.0000000<br>14.0000000 | 1.0000000<br>0 | 18.0000000<br>18.0000000 |

#### The MEANS Procedure

##### Gender=Female

| struggle with balancing work and childcare | N Obs | Variable | Label | N | Mean | Std Dev | 5th Pctl | 50th Pctl | 95th Pctl | Minimum | Maximum |
|---|---|---|---|---|---|---|---|---|---|---|---|
| no | 55 | AscoreR | HADS Anxiety score | 55 | 9.8000000 | 3.8412961 | 4.0000000 | 10.0000000 | 18.0000000 | 0 | 19.0000000 |
| | | DscoreR | HADS Depression score | 55 | 6.1454545 | 3.7683781 | 1.0000000 | 6.0000000 | 12.0000000 | 1.0000000 | 18.0000000 |
| yes | 53 | AscoreR | HADS Anxiety score | 53 | 10.0943396 | 3.9771663 | 5.0000000 | 10.0000000 | 18.0000000 | 1.0000000 | 20.0000000 |
| | | DscoreR | HADS Depression score | 53 | 6.5660377 | 2.8923602 | 2.0000000 | 6.0000000 | 12.0000000 | 2.0000000 | 14.0000000 |
| missing | 19 | AscoreR | HADS Anxiety score | 19 | 9.8421053 | 3.7603658 | 3.0000000 | 10.0000000 | 18.0000000 | 3.0000000 | 18.0000000 |
| | | DscoreR | HADS Depression score | 19 | 5.8421053 | 3.3708416 | 2.0000000 | 5.0000000 | 14.0000000 | 2.0000000 | 14.0000000 |

##### Gender=Male

| struggle with balancing work and childcare | N Obs | Variable | Label | N | Mean | Std Dev | 5th Pctl | 50th Pctl | 95th Pctl | Minimum | Maximum |
|---|---|---|---|---|---|---|---|---|---|---|---|
| no | 25 | AscoreR | HADS Anxiety score | 25 | 8.2000000 | 3.3541020 | 3.0000000 | 8.0000000 | 14.0000000 | 2.0000000 | 14.0000000 |
| | | DscoreR | HADS Depression score | 25 | 4.7600000 | 3.0177254 | 1.0000000 | 4.0000000 | 10.0000000 | 1.0000000 | 14.0000000 |
| yes | 16 | AscoreR | HADS Anxiety score | 16 | 10.1875000 | 3.6003472 | 4.0000000 | 11.0000000 | 16.0000000 | 4.0000000 | 16.0000000 |
| | | DscoreR | HADS Depression score | 16 | 6.3125000 | 3.6095937 | 1.0000000 | 5.5000000 | 14.0000000 | 1.0000000 | 14.0000000 |
| missing | 8 | AscoreR | HADS Anxiety score | 8 | 8.3750000 | 5.9506902 | 0 | 9.0000000 | 17.0000000 | 0 | 17.0000000 |
| | | DscoreR | HADS Depression score | 8 | 6.6250000 | 3.2043497 | 0 | 7.0000000 | 10.0000000 | 0 | 10.0000000 |
