## Supplemental Material 3 for "Symptoms of anxiety and depression in relation to work patterns during the first wave of the COVID-19 epidemic in Philadelphia PA: a cross-sectional survey"

**Supplemental Material 3: Principal Components Analysis of "worries" and HADS score for anxiety.**
**Eigenvectors**

| | Women | | | Men | | | All | | |
|---|---|---|---|---|---|---|---|---|---|
| **Prinipal component number** | **1** | **2** | **3** | **1** | **2** | **3** | **1** | **2** | **3** |
| Proportion correlation explained | 0.39 | 0.13 | 0.1 | 0.41 | 0.14 | 0.1 | 0.4 | 0.13 | 0.11 |
| HADS Anxiety score | **0.29** | 0.04 | **0.47** | **0.30** | 0.12 | **0.48** | **0.30** | 0.06 | **0.43** |
| I will be infected | **0.29** | **0.59** | -0.11 | **0.29** | **0.55** | -0.28 | **0.29** | **0.58** | -0.11 |
| I will infect my family | **0.24** | **0.68** | -0.06 | **0.23** | **0.65** | 0.26 | **0.24** | **0.68** | -0.01 |
| I will not be able to cope with the work | **0.33** | 0.00 | **0.22** | **0.39** | -0.04 | 0.01 | **0.35** | 0.00 | 0.19 |
| I will become poor | **0.39** | **-0.25** | **-0.24** | **0.38** | -0.19 | -0.19 | **0.38** | **-0.25** | **-0.26** |
| I will be short of food | **0.40** | **-0.27** | **-0.26** | **0.40** | -0.16 | **-0.31** | **0.40** | **-0.25** | **-0.28** |
| I will be short of medicines | **0.39** | -0.16 | **-0.32** | **0.39** | -0.09 | **-0.41** | **0.39** | -0.15 | **-0.35** |
| I will fail myself and my family | **0.39** | -0.12 | 0.01 | **0.34** | -0.19 | **0.50** | **0.37** | -0.13 | 0.07 |
| I will be confined at home and not able to leave | **0.21** | -0.13 | **0.70** | **0.25** | -0.39 | 0.26 | **0.23** | -0.19 | **0.70** |
