## Supplemental Material 4 for "Symptoms of anxiety and depression in relation to work patterns during the first wave of the COVID-19 epidemic in Philadelphia PA: a cross-sectional survey"

**Supplemental Material 4: Occupation of employment during epidemic and rate of anxiety and depression, along with relative rate (RR) and 95% confidence intervals (CI) of the difference in continuous HADS scores relative to sample mean symptoms scores.**

| Occupation | Women Total | Women Anxiety N, % | Women Anxiety RR# | Women Anxiety 95%CI | | Women Depression N, % | Women Depression RR## | Women Depression 95%CI | | Men Total | Men Anxiety N, % | Men Anxiety RR* | Men Anxiety 95%CI | | Men Depression N, % | Men Depression RR* | Men Depression 95%CI | |
|---|---|---|---|---|---|---|---|---|---|---|---|---|---|---|---|---|---|---|
| accountants, financial specialists, and other business operation | 33 | 12<br>36 | 0.92 | 0.80 | 1.07 | 2<br>6 | 0.95 | 0.78 | 1.16 | 13 | 2<br>15 | 0.81 | 0.61 | 1.06 | 0<br>0 | 0.78 | 0.55 | 1.10 |
| adminstrative support | 81 | 35<br>43 | 0.95 | 0.86 | 1.04 | 10<br>12 | 0.92 | 0.81 | 1.05 | 15 | 6<br>40 | 1.01 | 0.79 | 1.29 | 0<br>0 | 1.02 | 0.75 | 1.39 |
| architects and engineers | 10 | 4<br>40 | 0.93 | 0.72 | 1.19 | 2<br>20 | 1.13 | 0.81 | 1.57 | 15 | 4<br>27 | 0.90 | 0.70 | 1.16 | 1<br>7 | 0.88 | 0.64 | 1.20 |
| artists and performers | 19 | 10<br>53 | 1.11 | 0.93 | 1.32 | 4<br>21 | 1.22 | 0.96 | 1.55 | 8 | 6<br>75 | 1.28 | 0.94 | 1.75 | 2<br>25 | 1.35 | 0.92 | 2.00 |
| computer occupations | 17 | 10<br>59 | 1.04 | 0.86 | 1.26 | 2<br>12 | 0.89 | 0.68 | 1.16 | 14 | 4<br>29 | 0.86 | 0.66 | 1.12 | 0<br>0 | 0.77 | 0.55 | 1.09 |
| counselors, social workers, and community service occupations | 32 | 11<br>34 | 0.98 | 0.85 | 1.13 | 3<br>9 | 0.96 | 0.79 | 1.17 | 6 | 3<br>50 | 1.23 | 0.86 | 1.75 | 1<br>17 | 0.89 | 0.55 | 1.44 |
| healthcare practitioners and technicians | 55 | 22<br>40 | 0.96 | 0.86 | 1.07 | 8<br>15 | 0.99 | 0.85 | 1.15 | 19 | 7<br>37 | 1.10 | 0.89 | 1.37 | 2<br>11 | 1.08 | 0.82 | 1.43 |
| laywers and other legal occupations | 28 | 10<br>36 | 0.94 | 0.80 | 1.09 | 0<br>0 | 0.79## | 0.63 | 0.98 | 9 | 4<br>44 | 1.23 | 0.92 | 1.66 | 1<br>11 | 1.14 | 0.78 | 1.67 |
| librarians and curators | 16 | 7<br>44 | 1.03 | 0.85 | 1.26 | 3<br>19 | 1.07 | 0.82 | 1.39 | 2 | 1<br>50 | 1.17 | 0.63 | 2.16 | 0<br>0 | 1.15 | 0.53 | 2.52 |
| life, physical, and social scientists | 57 | 21<br>37 | 0.96 | 0.86 | 1.07 | 4<br>7 | 0.85 | 0.73 | 1.00 | 21 | 8<br>38 | 1.18 | 0.96 | 1.45 | 1<br>5 | 1.03 | 0.79 | 1.35 |
| managers | 115 | 60<br>52 | 1.09# | 1.00 | 1.18 | 17<br>15 | 1.04 | 0.93 | 1.16 | 47 | 12<br>26 | 0.95 | 0.81 | 1.11 | 4<br>9 | 0.90 | 0.73 | 1.09 |
| media and communication | 27 | 13<br>48 | 1.00 | 0.86 | 1.17 | 3<br>11 | 0.97 | 0.78 | 1.20 | 4 | 1<br>25 | 0.79 | 0.49 | 1.28 | 1<br>25 | 1.20 | 0.69 | 2.08 |
| other blue collar | 33 | 15<br>45 | 1.02 | 0.88 | 1.17 | 6<br>18 | 1.11 | 0.92 | 1.34 | 19 | 7<br>37 | 0.98 | 0.79 | 1.23 | 1<br>5 | 0.98 | 0.74 | 1.30 |
| retail and other sales related occupations | 17 | 13<br>76 | 1.15 | 0.96 | 1.39 | 4<br>24 | 1.20 | 0.93 | 1.55 | 7 | 1<br>14 | 0.82 | 0.57 | 1.18 | 1<br>14 | 1.29 | 0.85 | 1.96 |
| teachers | 90 | 45<br>50 | 1.02 | 0.93 | 1.11 | 13<br>14 | 1.00 | 0.88 | 1.13 | 25 | 4<br>16 | 0.81 | 0.66 | 1.00 | 3<br>12 | 0.92 | 0.72 | 1.19 |
| missing | 33 | 10<br>30 | N/A | | | 6<br>18 | N/A | | | 10 | 3<br>30 | N/A | | | 0<br>0 | N/A | | |

*none have p<0.05 after accounting for race, age, income, education, chidlren linig at home (not shown)

### after accounting for race, age, income, education, children living at home (not shown), evidence of association with managers increased to RR 1.10

#### after accounting for race, age, income, education, children living at home (not shown), evidence of association with lawyers and other legal occupations weakened to RR 0.80 (0.64, 1.00)
